# Non-Parametric Ancestry Adjustment for Polygenic Scores

**DOI:** 10.64898/2026.06.07.26355080

**Authors:** Daniel Mas Montserrat, Míriam Barrabés, Carlos D. Bustamante, Alexander G. Ioannidis

## Abstract

Modern polygenic risk scores (PRS) exhibit shifts correlated with ancestry, leading to erroneous predictions for non-European individuals when models are trained on predominantly European cohorts. Such shifts arise from, among other factors, (1) algorithmic limitations in the ability of PRS model training to detect causal variants, rather than nearby variants with ancestry-dependent correlations to the causal one, (2) under-representation of alleles with higher prevalence in non-European populations in the association study training, and (3) gene-by-environment interactions where the environment is correlated with genetic ancestry. Current ancestry-adjustment methodologies often discretize individuals into population categories and apply a simple affine mapping to reduce these genetic ancestry biases. However, such approaches provide suboptimal adjustments, particularly for admixed individuals. In this work, we introduce a detailed theoretical characterization of ancestry-dependent biases and propose novel methods based on non-parametric neighborhood techniques that provide more accurate empirical results and admit statistical consistency guarantees. Extensive experiments using the UK Biobank demonstrate the effectiveness of the proposed methods.

## 1. Introduction

Polygenic risk score (PRS) models (Box 1) predict susceptibility to complex diseases and heritable traits from genetic data by assuming a linear form in which each genomic position is assigned a weight. PRS have proven useful in areas ranging from genomic medicine to population-based screening. They have been widely applied to risk stratification for non-communicable chronic diseases, including cardiometabolic conditions such as type 1 and type 2 diabetes, dyslipidemia, and coronary artery disease, as well as multiple cancers, including breast, colorectal, prostate, ovarian, and lung cancers [14, 9]. Genomic risk estimates can support preventive care and enable more personalized treatments. PRS models are commonly derived from Genome-Wide Association Studies (GWAS), statistical learning approaches, or ensemble methods. GWAS-based PRS models process summary statistics using techniques like clumping to enforce sparsity and remove redundant genomic loci [16]. Statistical learning approaches fit linear models (e.g., LASSO) to estimate a weight for each variant [17]. Ensemble methods combine multiple PRS models derived from different heuristics and learning strategies [23].

The fact that PRS models aggregate contributions from many genomic positions, along with the central limit theorem (CLT), have together been a common starting point for theoretical formulations of polygenic scores under which a Gaussian distribution of predicted scores is typically assumed. Due to the nature of the algorithms used to construct these models, the predicted scores may exhibit arbitrary shifts and are often unitless (e.g., they do not correspond to logits or any calibrated scale). Therefore, they are typically normalized to follow a *z-score* distribution (mean 0, variance 1), allowing straightforward mapping to percentiles under the Gaussian assumption. Unfortunately, the shifts present in PRS models are often ancestry-dependent, with some population groups exhibiting larger deviations than others. If unaddressed, this artifact can lead to over- or under-estimated risk predictions for entire population groups. Such ancestry-dependent shifts originate in algorithmic limitations in model training, population imbalance, and, most importantly, mismatch between the training cohort and the target cohort. In particular, because most GWAS have been conducted in European-ancestry populations, PRS models derived from these studies—which constitute the majority of available PRS—often exhibit reduced transferability to non-European populations [12].

In order to remove ancestry-dependent biases, several adjustment strategies have been proposed, with two main approaches predominating: population discretization and linear adjustment. In the discretization approach, individuals are assigned to pre-defined ancestry categories, and the mean and standard deviation of the scores are computed within each category. When a new score is obtained, the individual is assigned to a corresponding ancestry category, and the mean and standard deviation are used to compute an ancestry-adjusted *z*-score. Alternatively, another common approach is to learn a linear mapping between an ancestry representation—typically the principal component analysis (PCA) projections of the genotypes—and the scores. This linear mapping is then used to residualize the scores, thereby removing ancestry-dependent biases. Both approaches have been adopted in research, clinical, and commercial applications; however, their underlying assumptions and limitations have not been explicitly analyzed, and more flexible adjustment strategies have not been extensively explored.

### Box 1

**Polygenic Risk Scores**

Polygenic risk scores (PRS) summarize an individual’s genetic liability by aggregating effects across many genetic variants, typically single-nucleotide polymorphisms (SNPs), which are genomic positions at which a single DNA base varies amongst individuals in the population. In autosomes (the majority of the genome), these SNPs consist of two alleles, one inherited from each parent. Let *S* denote the number of SNPs included in the score, and let *g* ∈ {0, 1, 2}^*S*^ denote the genotype sequence across these loci, where each entry *g*_*s*_ counts the number of copies (0, 1, or 2) of a designated effect allele at SNP *s*. In a standard formulation, the PRS is defined as a weighted sum of these allele counts,

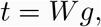

where *W* denotes the fitted PRS model (e.g., a vector of SNP effect sizes or, more generally, a linear operator) and *t* is the resulting scalar score. In parallel, genetic ancestry is often represented via an inferred variable obtained from an ancestry prediction function,

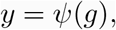

where *ψ* may be a linear mapping, such as a projection of the allele count vector onto principal component coordinates in PCA, or a non-linear mapping, as in local ancestry inference and related genetic clustering models. Because *t* aggregates contributions from many loci that are often treated as approximately weakly dependent, it is commonly modeled as approximately Gaussian by the central limit theorem, i.e., *t* ≈ *N* (*µ, σ*^2^) under suitable regularity conditions. For convenience, many analyses further assume that the PRS *t* is independent of ancestry *y*. However, in practice *t* and *y* are generally dependent due to population structure induced allele-frequency differences between populations and differential linkage disequilibrium patterns (allele correlations) across ancestries that change the effect sizes of non-causal alleles in the model that are correlated to the causal one. This simplifying assumption therefore fails in real applications and motivates ancestry adjustment techniques to reduce confounding and improve portability of PRS across diverse populations.

In this work, we develop a theoretical framework for understanding ancestry adjustment techniques for polygenic risk scores and introduce a new family of methods: non-parametric neighborhood-based adjustments. These techniques are designed to provide greater flexibility while making fewer assumptions about the underlying structure of ancestry-related biases. We present a theoretical characterization, together with experimental results, demonstrating that the proposed methods outperform traditional approaches.

## 2. Main Results

### 2.1. Affine Models

We assume a simple generative model for the raw polygenic risk score (PRS),

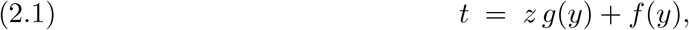

where *y* ∈ ℝ^*d*^ is a *d*-dimensional representation of ancestry (for example, a set of principal components), *t* is the observed (raw) PRS, and *z* is the latent “true” ancestry-free standardized score. The functions *f* (*y*) and *g*(*y*) *>* 0 capture systematic ancestry effects: *f* (*y*) represents an ancestry-dependent location (mean) shift, while *g*(*y*) represents an ancestry-dependent scaling (dispersion) factor. Throughout, we adopt two common assumptions: first, that the latent score is standard normal, *z* ∼ *N* (0, 1), and second, that it only captures polygenic factors, removing the signal from the ancestry representation. Namely, obtaining a score that is independent of the ancestry representation, *z* ⊥ *y*.

Under these assumptions, the conditional distribution of the raw score is Gaussian, *t* | *y* ∼ *N*, *f* (*y*), *g*^2^(*y*) so that its conditional mean and variance are E[*t* | *y*] = *f* (*y*), and Var(*t* | *y*) = *g*^2^(*y*), respectively. Thus, the observed *raw* score *t* is shifted by *f* (*y*) and scaled by *g*(*y*), with both location and scale depending on ancestry, making *t* dependent on *y*. Different ancestry adjustment techniques impose additional assumptions on the structure of *f, g*, and *y*. Current ancestry-adjustment techniques can be framed as learning functions *A*(*y*) and *B*(*y*) such that an ancestry-adjusted score is obtained via an affine transformation of the form:

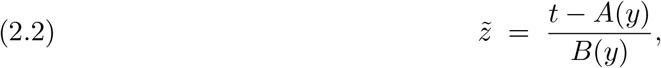

with the property that, for every ancestry configuration *y*, the adjusted score is centered and standardized, 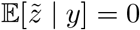, and 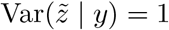. Imposing these conditional mean and variance constraints, the unique optimal affine standardization of *t* that achieves conditional mean 0 and conditional variance 1 for each ancestry *y* is given by *A*(*y*) = *f* (*y*) and *B*(*y*) = *g*(*y*) (see Appendix A.1). In practice, we observe *t* and *y*, but *g* and *f* are unknown. Given *t* and *y*, our goal is to construct an estimate 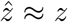 that approximates the true ancestry-independent risk score. The traditional ancestry-adjustment methods discussed below can be viewed as different approaches to estimating the nuisance functions 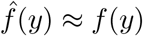 and *ĝ*(*y*) ≈ *g*(*y*) from data, after which the adjusted score is obtained by substituting these estimates into the standardization above.

### 2.2. Traditional ancestry-adjustment techniques

#### 2.2.1. Adjustment using ancestry categories

##### Model with ancestry categories

A widely adopted ancestry adjustment method is based on a categorical modeling of ancestry [14, 10]. The ancestry representation is partitioned into *K* discrete *ancestry categories A*_1_, …, *A*_*K*_, with *C*(*y*) ∈ {1, …, *K*} denoting the category label of an individual with ancestry descriptor *y*. The key modeling simplification is that ancestry-dependent location and scale are assumed to be constant within each category *k*. Specifically, for all *y* such that *C*(*y*) = *k*, we assume *f* (*y*) = *f*_*k*_ and *g*(*y*) = *g*_*k*_. Under the common assumptions, this implies that the conditional distribution of the raw PRS depends on *y* only through its category: 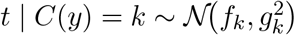.

##### Estimation within ancestry categories

Given observations (*t*_*i*_, *y*_*i*_) for *i* = 1, …, *n*, define for each category *k* the index set *I*_*k*_ = { *i* : *C*(*y*_*i*_) = *k* }, with sample size *n*_*k*_ = |*I*_*k*_|. Within category *k*, we estimate the category-specific mean by the sample average

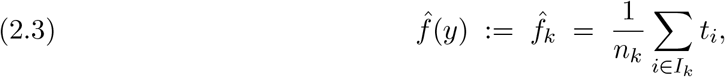

and the category-specific variance by the usual unbiased sample variance

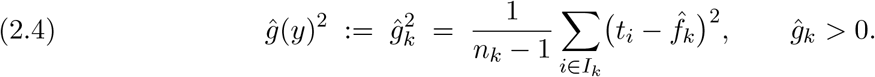

These estimates define piecewise-constant functions of ancestry through the mapping *C*(·), namely 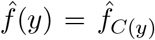 and *ĝ*(*y*) = *ĝ*_*C*(*y*)_ . The ancestry-adjusted score for individual *i* is then obtained via the affine standardization:

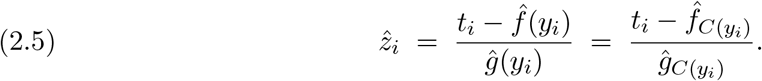

In the setting where *t*_*i*_ is generated according to this category-dependent model, 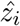 is consistent, with 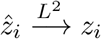. Proofs are provided in Appendix A.2.

#### 2.2.2. Linear ancestry adjustment

##### Mean-only linear model

A common parametric approach to ancestry adjustment assumes that the ancestry-dependent location varies linearly with the ancestry descriptor, while the scale is constant [20, 22]. Specifically, for an ancestry vector *y* ∈ ℝ^*p*^, we assume *f* (*y*) = *α*+*β*^⊤^*y* and *g*(*y*) = *σ >* 0, so that the raw PRS admits the Gaussian location-scale representation

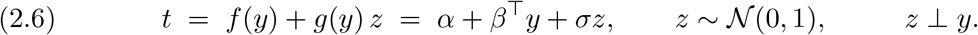

Under this model, the conditional distribution of the raw PRS is: *t* | *y* ∼ *N* *α* + *β*^⊤^*y, σ*^2^ . *Estimation under constant scale*. Given observations (*t*_*i*_, *y*_*i*_) for *i* = 1, …, *n*, we estimate the mean parameters (*α, β*) via ordinary least squares in the regression model, *t*_*i*_ = *α* + *β*^⊤^*y*_*i*_ + *ε*_*i*_, with *ε*_*i*_ ∼ *N* (0, *σ*^2^). Let 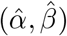 denote the OLS estimates, and define the fitted location function:

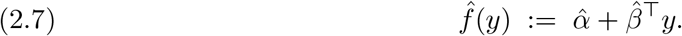

An unbiased estimator of the (constant) scale, based on the residuals *r*_*i*_, is given by

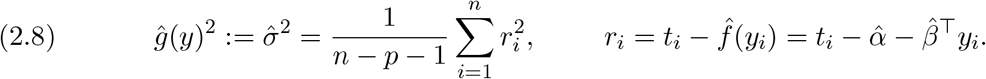

These estimates define the fitted location and scale functions 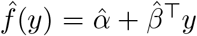 and 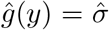. The ancestry-adjusted score for individual *i* is then obtained via the affine standardization:

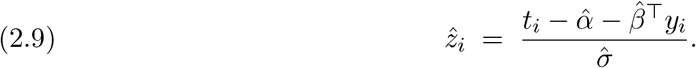

When *t*_*i*_ is generated according to this linear model, 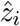 is asymptotically optimal, with 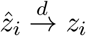. Proofs are provided in Appendix A.3.

Extensions of this approach that model the variance as a linear function of ancestry have been explored in [18, 6, 11]. Furthermore, techniques in [3] have combined the categorical and linear adjustment methods by fitting a linear model within each population group. We include all these techniques in our benchmarks.

#### 2.2.3. Traditional Model Limitations

The models described above impose strong assumptions on the form of *f* (*y*) and *g*(*y*), which may not accurately reflect the true structure of ancestry-dependent shifts. In particular, the discretization approach presents several limitations. First, it typically relies on pre-defined, self-identified labels (e.g., European, African, Asian) that may not align with the underlying genetic structure. Second, by treating each group as homogeneous, this approach ignores within-group genetic variation and genetic ancestry gradients between these “groups.” In particular it may fail to capture substantial within-ancestry diversity. Finally, it is ill-suited to admixed individuals with variable ancestry contributions from multiple self-identified groups. Similarly, the linear model assumption cannot capture non-linear relationships between ancestry and PRS shifts. In this work, we substantially relax these assumptions by requiring only Lipschitz continuity of *f* (·) and *g*(·), while providing asymptotically optimal estimators of the ancestry-adjusted score.

## 3. Non-parametric Neighborhood Models

### 3.1. Neighborhood Estimator

#### Affine Model

Our approach (Figure 1) builds on the same affine generative model introduced in (2.1), in which the observed *raw* (unadjusted) polygenic score *t* is expressed as a location–scale transformation of an unobserved ancestry-independent latent score *z*, i.e., *t* = *z g*(*y*) + *f* (*y*), where *y* ∈ ℝ^*d*^, *z* ∼ *N* (0, 1), and *z* ⊥ *y*. The functions *f* and *g* are assumed to be *L*_*f*_ - and *L*_*g*_-Lipschitz, respectively, with regard to the Euclidean (*ℓ*_2_) distance. In addition, *g* is assumed to be strictly positive and bounded away from zero, i.e., there exists *g*_*ℓ*_ *>* 0 such that *g*(*y*) ≥ *g*_*ℓ*_ for all *y*.

**Figure 1.**
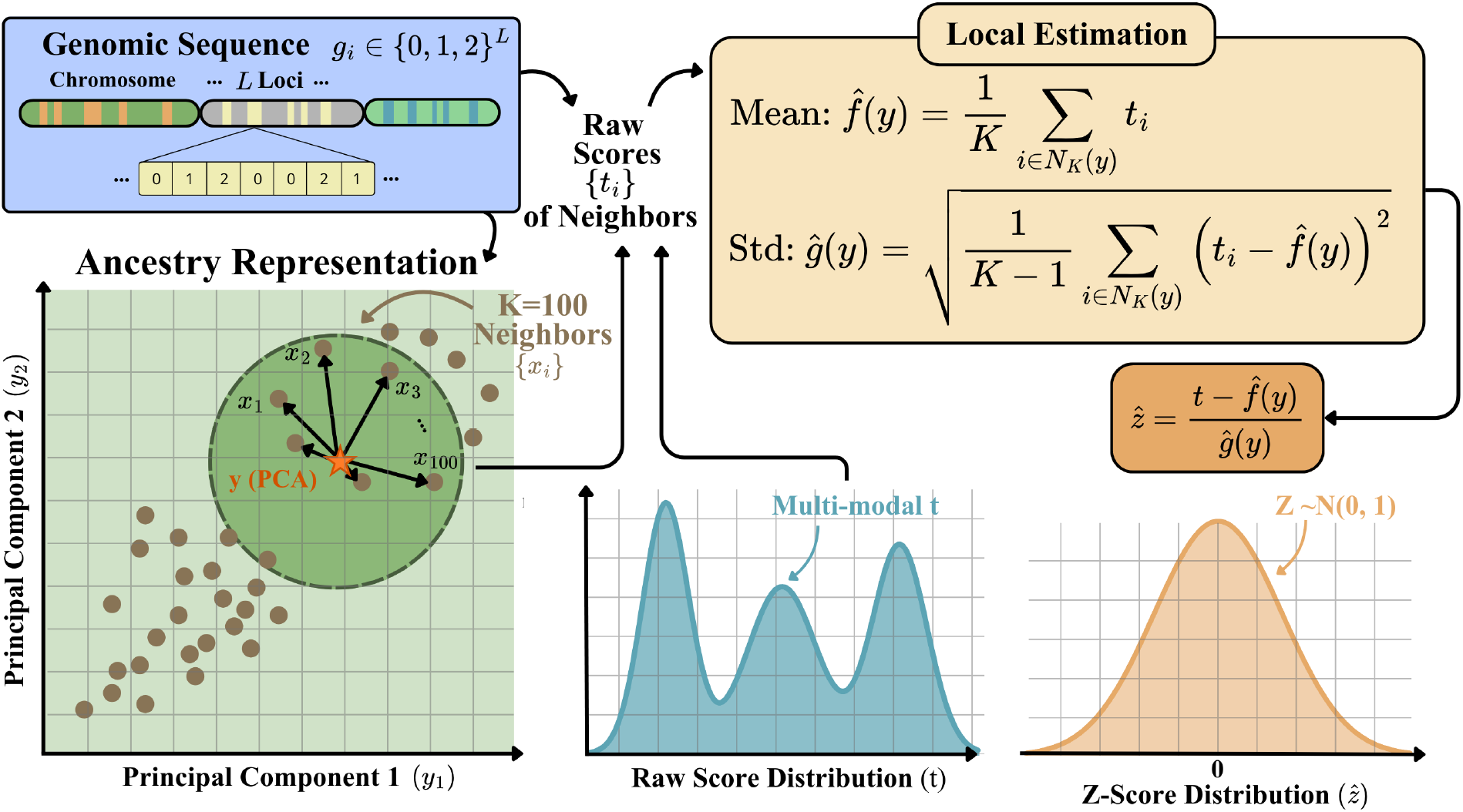
Neighborhood-based ancestry adjustment. Starting from a genomic sequence g_i_ ∈ {0, 1, 2}^L^, an ancestry representation y ∈ ℝ^d^ is inferred. For a target individual, the K-nearest neighbors {x_i_} are identified in the ancestry space using a distance metric. The corresponding raw polygenic scores {t_i_} from the neighbors are used to estimate the ancestry-dependent local location 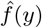 and scale ĝ(y). The adjusted score is then computed as 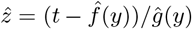. While the marginal distribution of raw scores t may be multi-modal due to population structure, the adjusted scores 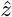 approximate a standard normal distribution.

#### Database of raw scores and K-neighborhoods

We are given a database of *M* score–ancestry pairs, *D*_*M*_ = {(*t*_1_, *x*_1_), (*t*_2_, *x*_2_), …, (*t*_*M*_, *x*_*M*_)}, where each observation follows the model *t*_*j*_ = *z*_*j*_ *g*(*x*_*j*_) + *f* (*x*_*j*_) for *j* = 1, …, *M*. The database *D*_*M*_ is assumed to be sufficiently large and diverse, covering a broad range of ancestries, so that local neighborhoods in genetic ancestry space can be used to accurately estimate ancestry-dependent effects.

Given a new individual with ancestry representation *y*, and a distance metric *d*(·, ·) on the genetic ancestry space, we define its *K*-nearest neighborhood of *y* as:

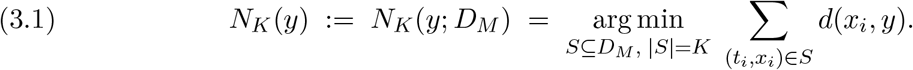

That is, *N*_*K*_(*y*; *D*_*M*_) consists of the *K* samples in *D*_*M*_ whose ancestry representations are closest to *y* under the metric *d*. A convenient representation of *N*_*K*_(*y*; *D*_*M*_) can be obtained by sorting the samples according to their distance to *y*. Let {*i*_(1)_, …, *i*_(*M*)_} be a permutation of {1, …, *M* } such that

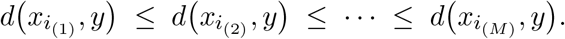

Then,

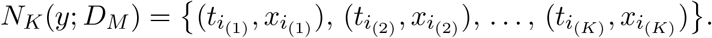

The two definitions are equivalent whenever the distances *d*(*x*_*i*_, *y*) are distinct; ties at the *K*-th position can be broken arbitrarily. For notational simplicity, we relabel indices so that *i*_(*k*)_ = *k* for *k* = 1, …, *K*.

#### *Estimator of* 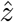

Given a *raw* score *t* with ancestry descriptor *y*, and its *K*-neighborhood *N*_*K*_(*y*; *D*_*M*_), we estimate the local ancestry-dependent location and scale by computing empirical statistics over the neighborhood. The local mean estimator is defined as:

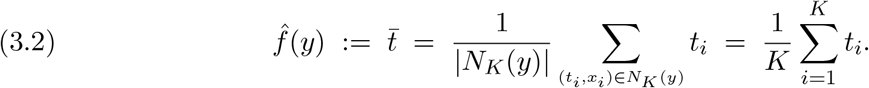

To estimate the local scale, we use a regularized version of the empirical standard deviation to ensure numerical stability and strict positivity. Specifically, for a fixed *ϵ >* 0, we define:

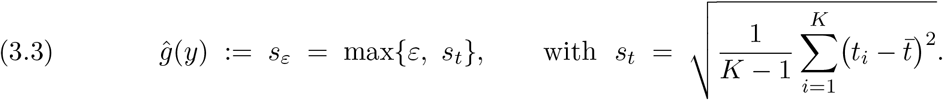

We assume that *ε* is chosen sufficiently small so that *ε < g*_*ℓ*_. The resulting estimator of the ancestry-adjusted score is then given by:

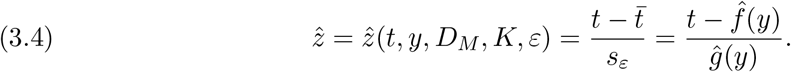

Intuitively, 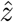 computes a *z*-score using the mean and standard deviation estimated from individuals with similar ancestry. Thus, the adjusted score reflects the relative genetic risk with respect to the *K*-nearest neighbors in ancestry space.

### 3.2. Mean Squared Error and Asymptotic Optimality

The expected mean squared error (MSE) of the estimator defined in Equation (3.4) admits an upper bound that decomposes into a variance term and a bias term. The variance decreases with the number of neighbors *K*, while the bias increases with *K*, reflecting the classical bias–variance trade-off in non-parametric estimation.

#### Proposition 3.1 (Quadratic bound with *K/M* bias control)

*Assume d* ≥ 2, *M* ≥ *K* ≥ 10, 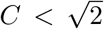, *and τ* ∈ (0, 1). *Let the ancestry representation y* ∼ *p be sampled independently of the reference set* 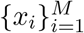, *where the density p is supported on a compact set S* ⊂ ℝ^*d*^ *with nonempty interior and finite diameter. Assume that p is bounded away from zero and infinity on S, i.e*., *there exist constants* 0 *< m* ≤ *M*_0_ *<* ∞ *such that m* ≤ *p*(*x*) ≤ *M*_0_ *for all x* ∈ *S. Moreover, assume that S satisfies the uniform interior-volume condition from Section C.3. Then the estimator* 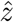 *satisfies*

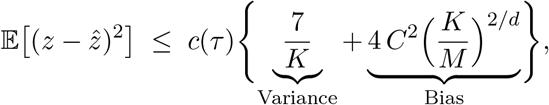

*where τ* := *ε/g*_*ℓ*_ ∈ (0, 1), *c*(*τ*) := 79*/*(*τ* ^2^(1 + *τ*)^2^), *and C* := *LC*_*p*_*/g*_*ℓ*_, *with L* := max{*L*_*f*_, *L*_*g*_} *denoting the maximum Lipschitz constant of f and g, and C*_*p*_ *denoting a distribution-dependent constant described in Corollary C.11*.

*Proof*. See Appendices B, C, and D.

We denote the upper bound in Proposition 3.1 by 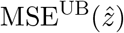. This bound provides insight into the behavior of the estimator through its explicit dependence on the parameters *K, M*, and *τ* = *ε/g*_*ℓ*_. First, the factor *c*(*τ*) depends on the ratio *τ* = *ε/g*_*ℓ*_. As *τ* → 1^−^, the factor *c*(*τ*) decreases and reaches its minimum at *τ* = 1. However, the bound is only valid for *τ <* 1, which is equivalent to requiring *ε < g*_*ℓ*_. This creates a trade-off: choosing *ε* closer to *g*_*ℓ*_ tightens the bound, but *ε* must remain strictly smaller than *g*_*ℓ*_ to preserve the validity of the bound and the associated consistency guarantees. Second, the bound shows that, under the stated assumptions, the estimator is consistent provided that the number of neighbors *K* grows sublinearly with respect to *M* . In this regime, both the variance term and the bias term vanish as *M* → ∞. This is formalized in the following proposition.

#### Proposition 3.2 (Consistency with 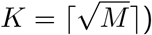

*Under the assumptions of Proposition 3.1, fix* 2 ≤ *d <* ∞, *let M* ≥ 100, *and define* 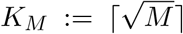. *Let* 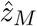 *denote the estimator constructed from M reference samples using K*_*M*_ *neighbors. Then*

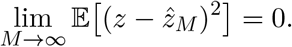

*Proof*. For *M* ≥ 100, we have 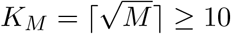 and *K*_*M*_ ≤ *M*, so Proposition 3.1 applies and gives

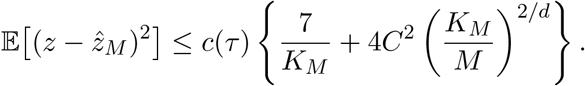

Moreover, since 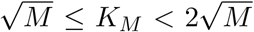, it follows that 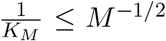 and 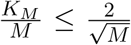. Substituting these bounds yields

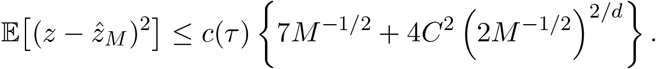

Since *d <*∞, we have 2*/d >* 0. Therefore, as *M* → ∞, we get that *M* ^−1*/*2^ → 0, and (2*M* ^−1*/*2^)^2/d^ → 0, which implies

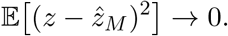

Proposition C.11 shows that the performance of the estimator can be characterized by the single constant *C*, which serves as a measure of the complexity of the ancestry-dependent distortion. When *C* = 0, the problem simplifies to a setting where the distortion of the raw score *t* is no longer ancestry-dependent. Moreover, as established in the proposition below, *C* is lower bounded by the Pearson correlation *ρ*(*t, y*) between the raw score *t* and the ancestry descriptor *y* (see Box 2). A larger value of *ρ*(*t, y*) indicates that *t* carries a stronger ancestry signal, and thus that a more complex adjustment may be required. In contrast, *ρ*(*t, y*) = 0 suggests that *f* and *g* are constant and ancestry-independent. At the other extreme, when *ρ*(*t, y*) = 1, we have 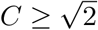 in this case, the raw score *t* captures only ancestry information, and there is no ancestry-independent score *z* to recover, rendering the bound invalid.

#### Box 2

**Pearson Correlation and Correlation Ratio**

Let *v* be a real-valued random variable with Var(*v*) ∈ (0, ∞), and let *w* ∈ ℝ^*d*^ be a random vector. Define the Pearson correlation as the maximal linear correlation between *v* and *w* by

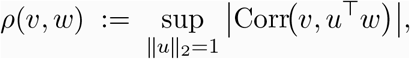

and define the correlation ratio of *v* on *w* by

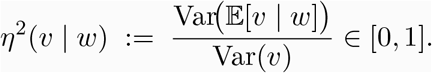

Then one always has the inequality *ρ*(*v, w*) ≤ *η*(*v* | *w*), which follows from the law of total covariance and the Cauchy-Schwarz inequality: for any ∥*u*∥_2_ = 1, letting *s* := *u* ^⊤^*w*, we have Cov(*v, s*) = Cov(E[*v* | *w*], *s*), and hence 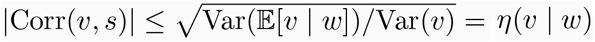, taking the supremum over *u* yields the claim.

#### Proposition 3.3 Bound on *ρ, η*, and *C*

*Let y* ∼ *p and y* ∈ ℝ^*d*^ *with covariance matrix* Σ := Cov(*y*), *C*_*p*_ *is a constant particular to the distribution of the ancestry representation p as described in Appendix C.3 with* 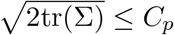 *and C* := *LC*_*p*_*/g*_*ℓ*_, *then*

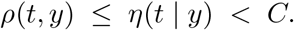

*Proof*. As noted above, *ρ*(*t, y*) ≤ *η*(*t* | *y*), so it suffices to bound *η*(*t* | *y*). Since *z* is independent of *y* and E[*z*] = 0, we have E[*t* | *y*] = *f* (*y*), and hence Var(E[*t* | *y*]) = Var(*f* (*y*)). Moreover, Var(*t* | *y*) = Var (*z g*(*y*) | *y*) = *g*(*y*)^2^. By the law of total variance, Var(*t*) = Var E[*t* | *y*] + E Var(*t* | *y*) = Var(*f* (*y*)) + E [*g*(*y*)^2^]. Substituting into the definition of *η* yields *η*^2^(*t* | *y*) = Var(*f* (*y*))*/* Var(*f* (*y*)) + E[*g*(*y*)^2^], and therefore

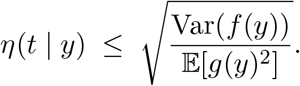

We next bound Var(*f* (*y*)). Since *f* is *L*_f_ -Lipschitz and *L* = max (*L*_*f*_, *L*_*g*_), we have |*f* (*y*) − *f* (E*y*)| ≤ *L*∥*y* − E*y*∥_2_. Thus, 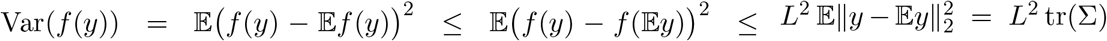. Since *g*(*y*) ≥ *g*_*ℓ*_, we have 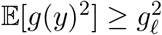. Combining these bounds,

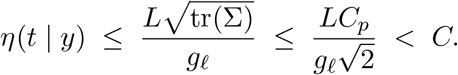

As *d* and *M* are fixed, and *ε* must be chosen sufficiently small to ensure consistency of the estimator, the only hyperparameter of 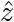 is the number of neighbors *K*. Proposition 3.1 can then be used to determine the optimal value *K*^⋆^ that minimizes the upper bound, as formalized in Proposition 3.4.

#### Proposition 3.4 (Optimal *K* minimizing the upper bound)

*Under the conditions of Proposition 3.1, consider the upper bound c*(*τ*){7*/K* + 4*C*^2^(*K/M*)^2*/d*^} *as a function of K* ∈ (0, ∞) *(treating K as continuous). The unique minimizer is*

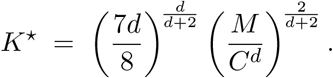

*In practice, one may take K* = ⌈*K*^⋆^⌉ *(and enforce* 10 ≤ *K* ≤ *M)*.

*Proof*. Ignoring the positive prefactor *c*(*τ*), we minimize *ϕ*(*K*) := 7*K*^−1^ + 4*C*^2^*M* ^−2*/d*^*K*^2*/d*^ over *K >* 0. Differentiating yields 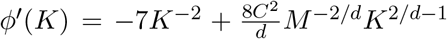. Thus, *ϕ*^′^(*K*) = 0 if and only if 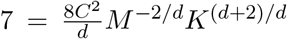, i.e., 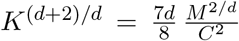 . Solving yields the stated expression for *K*^⋆^. Since 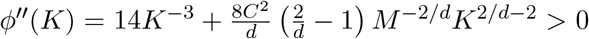 0 for all *K >* 0 and *d* ≥ 2, this critical point is the unique minimizer.

#### Corollary 3.5 Optimal *K* when *d* = 2

*In the setting of Proposition 3.4 with d* = 2, *the minimizer simplifies to*

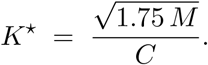

*Moreover, if ρ*(*t, y*) ≤ *C, then*

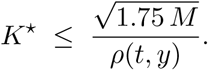

The scaling 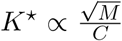 is consistent with the requirement that *K* grows sublinearly with respect to *M*, and provides an intuitive interpretation of *K*-NN. Specifically, small values of *K* lead to more complex decision boundaries in *K*-NN regressors and classifiers, whereas larger values of *K* produce smoother functions. This behavior is reflected in the inverse relationship between the number of neighbors *K* and the complexity constant *C*.

## 4. Bounds on the MSE from Moments and Correlation with an Independent Covariate

### Evaluating Adjusted Scores

Given a generic score estimator 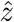 produced by an ancestry adjustment technique, it is difficult to properly assess its quality, since the true adjusted score *z* is unobservable. Moreover, the underlying distortion functions *f* and *g* are unknown, and only *t* and *y* are accessible. Simply measuring the Gaussianity of 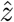 fails to capture potential residual dependence between 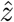 and *y*. Therefore, a metric that can be empirically approximated and quantifies the deviation of 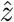 is from the true risk score *z* is required. Below, we propose two such metrics that provide upper and lower bounds on the true MSE.

Let *z* ∼ *N* (0, 1), and let 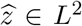 be a real-valued random variable with mean 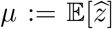 and variance 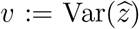. Let *y* ∈ ℝ^*d*^ have finite second moments and be independent of *z*. Define 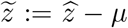, so that 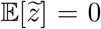 and 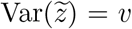. Set 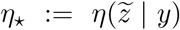 and 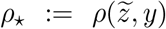, where *η*(· | ·) and *ρ*(·, ·) are defined in Box 2.

#### Proposition 4.1 Two-sided MSE bounds from an independent covariate

*With the setup above*,

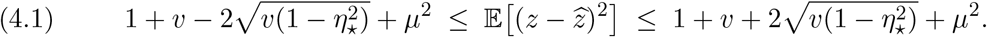

*Moreover, ρ*_⋆_ ≤ *η*_⋆_ *by Box 2, and hence* (4.1) *also holds with η*_⋆_ *replaced by ρ*_⋆_. *In particular, setting η*_⋆_ = 0 *(equivalently, ρ*_⋆_ = 0*) yields*

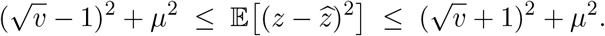

*Proof*. Expanding with 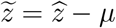 and using Var(*z*) = 1 gives

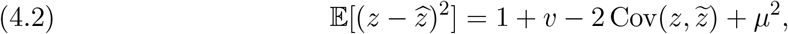

so it suffices to bound 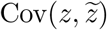.

Let 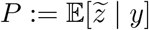S and 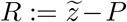 . Since *y* ⊥ *z*, any measurable function of *y* is independent of *z*. Hence, Co v(*z, P*) = 0, and thus 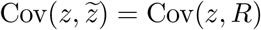. By the law of total variance and the definition of *η*_⋆_ (Box 2),

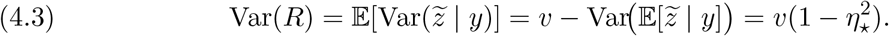

By the Cauchy–Schwarz inequality,

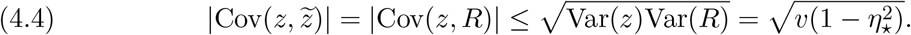

Substituting 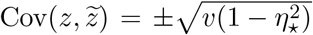 into (4.2) yields (4.1). The *ρ*_⋆_ specialization follows from *ρ*_⋆_ ≤ *η*_⋆_ in Box 2.

We refer to the lower bound in Equation (4.1) as 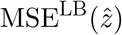.

## 5. Non-parametric Kernel Models

### Weighted estimator for 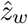

Given a *raw* score *t* with ancestry descriptor *y* and its *M* - neighborhood 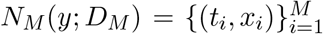 (with *d*(*x*_*i*−1_, *y*) ≤ *d*(*x*_*i*_, *y*)), let *w* = (*w*_1_, …, *w*_*M*_) be nonnegative weights such that 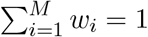. Define the weighted empirical mean

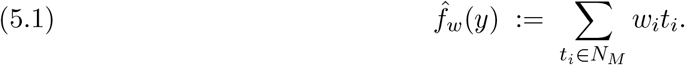

To define a weighted analogue of the sample standard deviation—recovering to the usual *K*-NN standard deviation when *w* places uniform mass on the closest *K* neighbors—we set

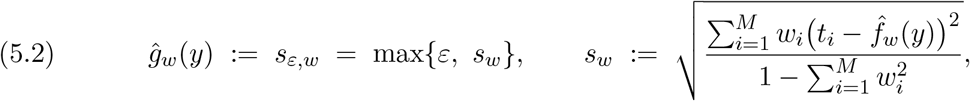

where we assume 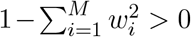 (i.e., the weights are non-degenerate). Assuming *ε* is chosen sufficiently small (e.g., *ε < g*_ℓ_), the weighted ancestry-adjusted score is

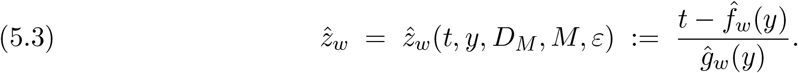

The original *K*-NN estimator 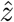 is recovered by choosing weights that assign uniform mass to the closest *K* neighbors and zero elsewhere:

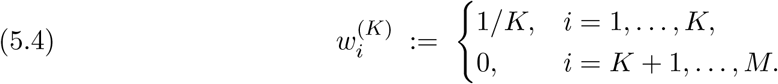

Indeed, 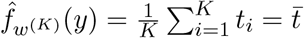 and 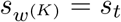, hence 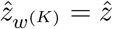.

### Risk-optimal weighting

Let *z* denote the target ancestry-adjusted score, and define the mean-squared error

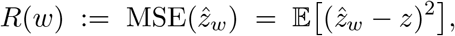

where the expectation is taken over the randomness in the data (and neighborhood construction). Define

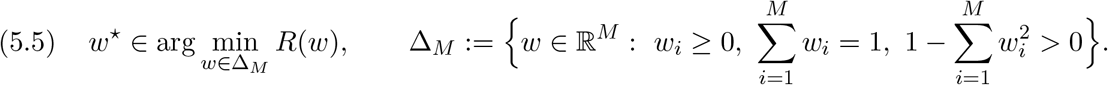

#### Proposition 5.1 (No-worse MSE than the *K*-NN estimator)

*Fix any* 1 ≤ *K* ≤ *M, and let* 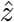 *denote the corresponding K-NN estimator in* (3.4). *Then*

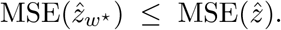

*Proof*. For any fixed *K*, the weight vector *w*^(*K*)^ defined in (5.4) is feasible, i.e., *w*^(*K*)^ ∈ Δ_*M*_, and satisfies 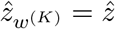. Therefore,

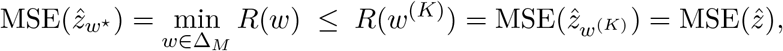

which proves the claim.

### Kernel-induced weights

A convenient and widely used specialization is to define the weights as a normalized kernel function of the ancestry distance between the query ancestry descriptor *y* and each neighbor descriptor *x*_*i*_. Let *d*(·, ·) denote the ancestry-space metric used to construct neighborhoods, and let *ω* : ℝ_≥0_ → ℝ_≥0_ be a nonnegative kernel profile. For a bandwidth (scale) parameter *h >* 0, define the unnormalized kernel weights

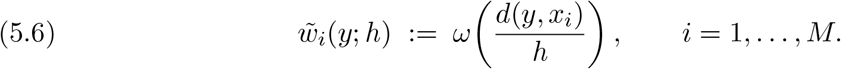

These are normalized to sum to one:

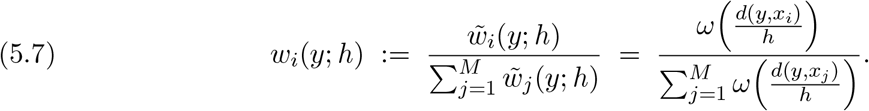

This construction ensures *w*_*i*_(*y*; *h*) ≥ 0 and 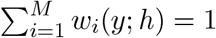, so that *w*(*y*; *h*) ∈ Δ_*M*_ provided the denominator is nonzero and the weights are non-degenerate (i.e., 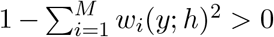). In practice, to improve stability in regions with sparse neighbors, the kernel values for the *q* closest samples may be clamped to a constant. In this work, we use *q* = 50 and clamp these values to the minimum kernel value among the *q* nearest neighbors. Substituting *w*(*y*; *h*) into (5.1)–(5.3) yields the kernel-weighted ancestry-adjusted score

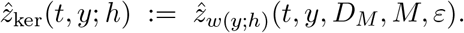

Common choices include compactly supported kernels (e.g., Epanechnikov, tri-cube) and exponentially decaying kernels (e.g., Gaussian, Laplacian), which place greater emphasis on closer ancestries. The bandwidth *h* controls the effective number of neighbors receiving non-negligible weight and can be selected via cross-validation or other data-driven procedures. Finally, note that the uniform *K*-NN estimator is recovered as a special case by using a *box* kernel supported on the *K* nearest neighbors. Equivalently, this corresponds to setting *ω*(*u*) = I{*u* ≤ 1} and choosing *h* such that exactly *K* points satisfy *d*(*y, x*_*i*_) ≤ *h*), which induces weights *w*_*i*_ = 1*/K* for those *K* points and *w*_*i*_ = 0 otherwise.

### Joint risk optimization over kernel choice and bandwidth

The kernel-weight construction (5.7) depends on both the bandwidth *h* and the kernel profile *ω*. Let Ω denote an admissible class of nonnegative kernel profiles (e.g., a finite set including box, Epanechnikov, tri-cube, Gaussian, and Laplacian kernels), and let ℋ ⊂ (0, ∞) denote an admissible set of bandwidths chosen so that *w*(*y*; *ω, h*) ∈ Δ_*M*_ (i.e., normalization is well-defined and weights are non-degenerate). Define the joint kernel risk

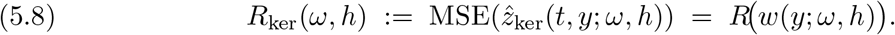

A *risk-optimal kernel-bandwidth pair* is any solution

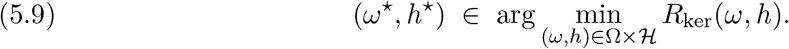

When Ω is finite, the optimization in (5.9) reduces to selecting the best kernel by minimizing over *h* for each candidate:

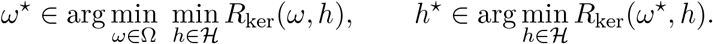

In practice *R*_ker_(*ω, h*) is unknown and must be approximated using an empirical risk estimate (e.g., an empirical estimate of MSE^LB^), yielding data-driven selections 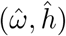. This joint tuning makes the bias–variance trade-off explicit not only through the scale *h*, but also through the tail behavior and support of *ω*: compactly supported kernels enforce hard locality, while exponentially decaying kernels smoothly downweight distant ancestries, potentially exhibiting different robustness to heterogeneity and outliers in the neighborhood.

## 6. Experimental Results

### Theoretical bounds under simulations

We construct a simulation environment to assess the validity of the proposed theoretical bounds. Namely, we sample the latent ancestry-independent score *z* ∼ *N* (0, 1), and generate ancestry representations *x*_*i*_, for *i* = 1, …, *M*, uniformly in a *d*-dimensional ball of radius *R*. As distortion functions, we use *f* (*x*) = *L*_*f*_ ||*x* − *R***1**|| and *g*(*x*) = *L*_*g*_||*x* − *R***1**|| + *g*_ℓ_, so that both the shift and the scale vary linearly with the distance of *x* from the reference point *R***1**. To emulate a worst-case distortion, we sample the test set from points surrounding *R***1**. The simulation parameters are *L*_*f*_ = *L*_*g*_ = 0.5, *R* = 1, *d* = 2, *g*_ℓ_ = 5, and *M* = 45000, with a numerically estimated constant *C*_*p*_ = 1.8. To ensure that empirical errors closely approximate their expectation, we use a large test set of size *D* = 30000 and average results over 60 independent repetitions. Figure 5 compares empirical performance with theoretical predictions as a function of the number of neighbors *K*. The black curve represents the empirical mean squared error 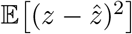, which exhibits the characteristic U-shaped behavior. The orange curve corresponds to the theoretical upper bound MSE^UB^ (Proposition 3.1), while its decomposition into bias and variance components is shown by the dashed blue curves: the darker blue dashed line represents the bias term, which decreases with smaller *K*, and the lighter blue dashed line represents the variance term, which decreases with larger *K*. Their sum yields the theoretical upper bound and highlights the bias–variance trade-off. The two-sided bounds from Equation (4.1) are shown as the lower bound (dark green) and the corresponding upper envelope (light green), which bracket the empirical error. Notably, the lower bound provides a very tight approximation of the empirical MSE, acting as a good evaluation metric when the true ancestry-independent score *z* is unavailable. The dashed olive vertical line indicates the optimal *K*^⋆^ predicted by the theoretical upper bound, which closely aligns with the empirically observed optimum. However, since the upper bound MSE^UB^ is derived under worst-case conditions induced by the Lipschitz assumption, it does not necessarily provide accurate estimates of the optimal *K*^⋆^ unless the underlying *f, g*, and *y* satisfy such worst-case behavior, as enforced in this simulation.

### Empirical Analysis with the UK Biobank

We conduct our empirical analysis on the UK Biobank [4], a large-scale dataset containing numerous genetic sequences from individuals in the UK. We select a subset including all individuals with non-European self-reported ethnicity, together with a subset of individuals of European descent to obtain a diverse and balanced dataset. After filtering, the dataset contains a total of 64000 individuals. Genotype sequences are processed using the `snputils` library [2]. We evaluate a total of 4000 PRS models from the PGSCatalog, covering a wide range of traits, including cancer, cardiometabolic diseases, and psychological traits, among others. Predictions for all models are obtained using the `StrataRisk™` pipeline [13, 9]. Models producing highly unstable predictions are filtered out.

We represent ancestry using a Principal Component (PC) model trained on all autosomal chromosomes from the 1000 Genomes Project [21]. As is common in practice, we retain up to 10 PC dimensions. Figure 3 shows the first two components, colored by self-reported ethnicity. Individuals with unknown or unspecified ethnicity are excluded from the visualization. The resulting ancestry representation exhibits an approximate convex hull structure, with European, African, and Asian ancestries located near the vertices, and admixed individuals lying in the interior. Similar structure is observed with alternative representations, including archetypal analysis [7] and other probabilistic representations [5, 19].

**Figure 2.**
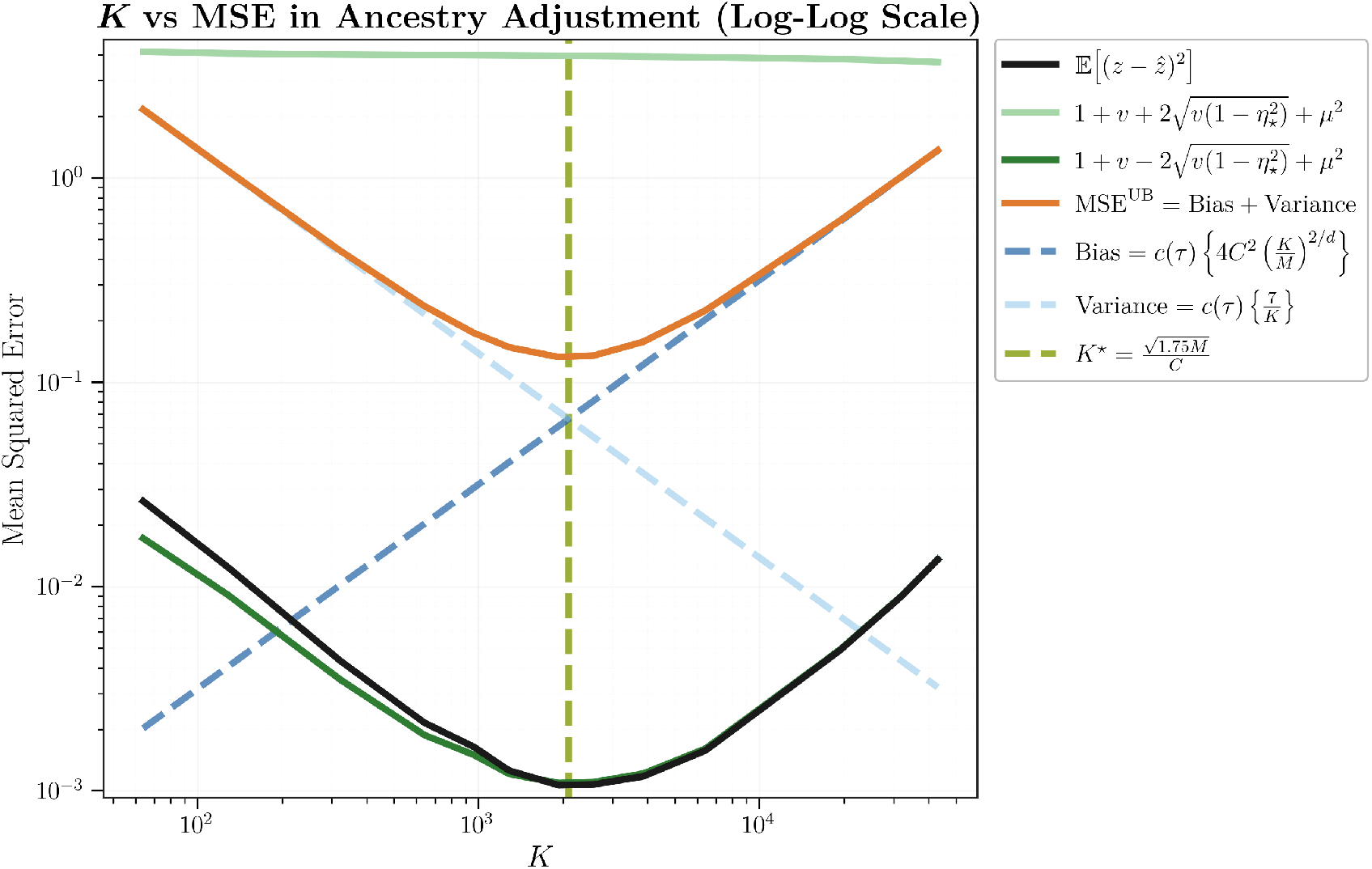
Theoretical bounds under simulation. Empirical mean squared error (MSE) of the neighborhood-based estimator as a function of the number of neighbors K (black), together with the empirical lower and upper bounds from Equation (4.1), shown as the lower bound (dark green) and the corresponding upper envelope (light green), computed from the correlation ratio, mean, and variance of the adjusted score, and the theoretical upper bound from Proposition 3.1 (orange), which depends on K, the reference set size M, the complexity constant C, and the ancestry dimension d. The latter is decomposed into bias and variance components (blue dashed lines), illustrating the bias–variance trade-off. The dashed olive vertical line marks the optimal K^⋆^ predicted by the theoretical upper bound. Results are averaged over 60 repetitions using a test set of size D = 30000. The simulated setting uses L_f_ = L_g_ = 0.5, R = 1, d = 2, g_ℓ_ = 5, M = 45000, and C_p_ = 1.8. The figure shows that the empirical MSE is tightly approximated by the lower bound, while the theoretical upper bound captures the bias-variance trade-off and predicts an approximately correct optimal value of K.

**Figure 3.**
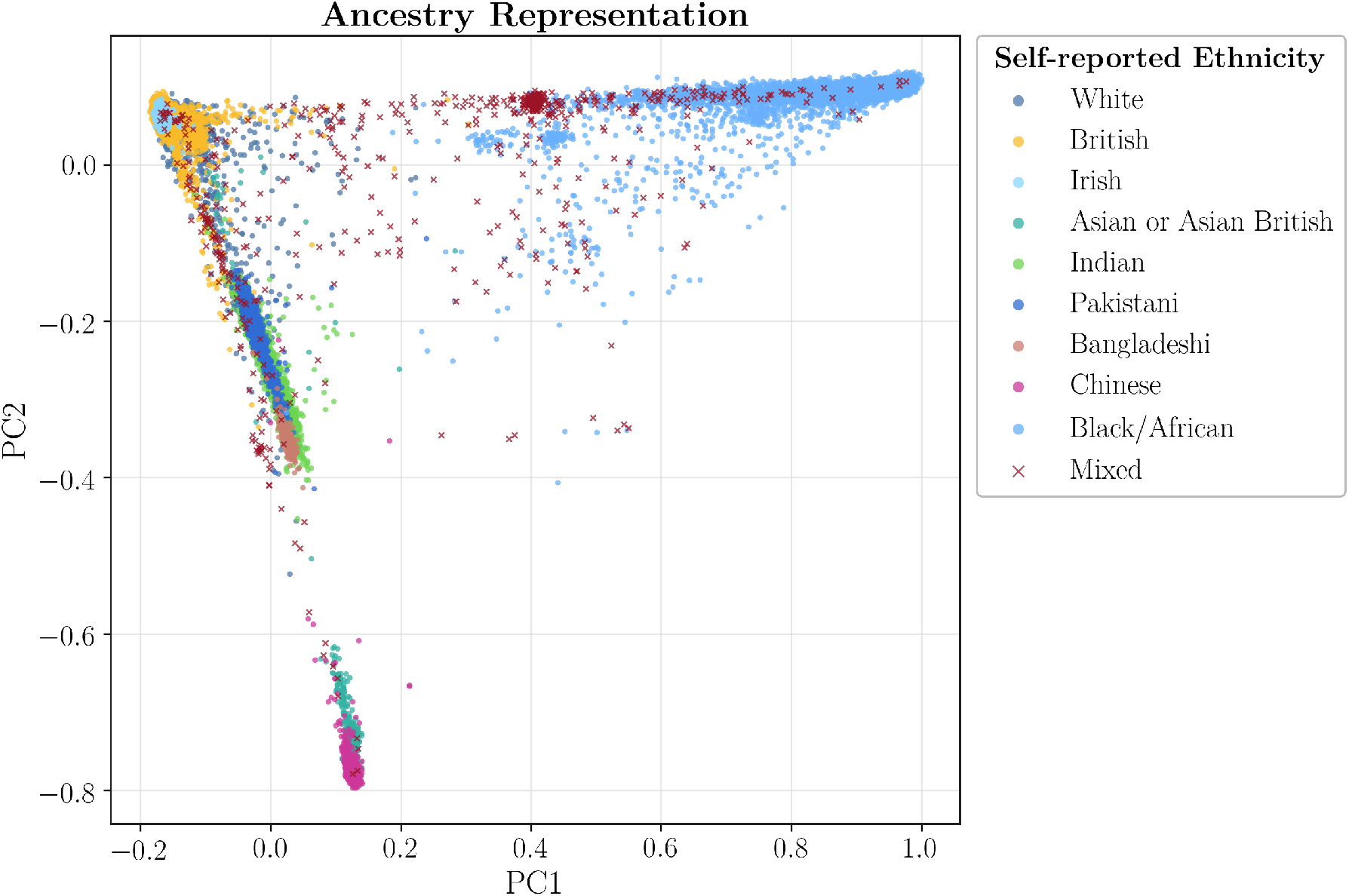
PCA of ancestry representation in the UK Biobank. Scatter plot of the first two principal components (PC1 and PC2) of the inferred genetic ancestry representation for UK Biobank participants, where each point corresponds to an individual and is colored by self-reported ethnicity. The projection reveals clear structure across major ancestry groups, with substantial clustering for White, British, Irish, South Asian, East Asian, and Black/African individuals, while individuals reporting Mixed ethnicity are more broadly distributed across the embedding. Overall, the figure illustrates that the learned ancestry representation captures meaningful population structure and aligns well with self-reported ethnicity, while also reflecting overlap and admixture between groups.

In all experiments, we use leave-one-out (LOO) estimators, where each sample *i* is evaluated using a model trained on the full dataset excluding that sample. All our empirical predictors admit efficient LOO implementations, avoiding the need to fit 64000 models. This approach enables the use of the full dataset for both training and evaluation while avoiding over-fitted estimates. The same LOO approach is used to estimate 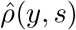, where *s* denotes either the predicted 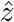 or the raw score *t*, as well as 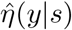. To approximate 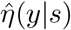 efficiently while providing a fast LOO estimator, we employ a random ReLU feature expansion *ϕ* [8] of *y*, and compute 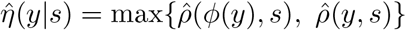.

### Qualitative Analysis

To provide intuition about ancestry-dependent biases in real PRS models, we visualize the estimated functions 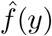 and 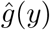 for a representative model, together with the distributions of the raw score *t* (pre-adjustment), and the adjusted score 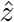 (post-adjustment). Figure 4 shows that the raw PRS score *t* (PGS001937) exhibits a highly multimodal distribution, while the transformed score 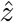 is substantially closer to Gaussian. We observe that the learned affine shift 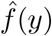 induces a distortion approximately proportional to the distance from the European population. This is aligned with the fact that most models are trained predominantly on European-descent samples, leading to out-of-distribution shifts when applied to non-European populations. However, non-linear behavior can also be observed, highlighting the importance of using flexible models. Similarly, 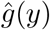 shows a significant increase in the scaling factor for individuals with admixed European and African ancestry, while exhibiting a highly non-linear mapping.

**Figure 4.**
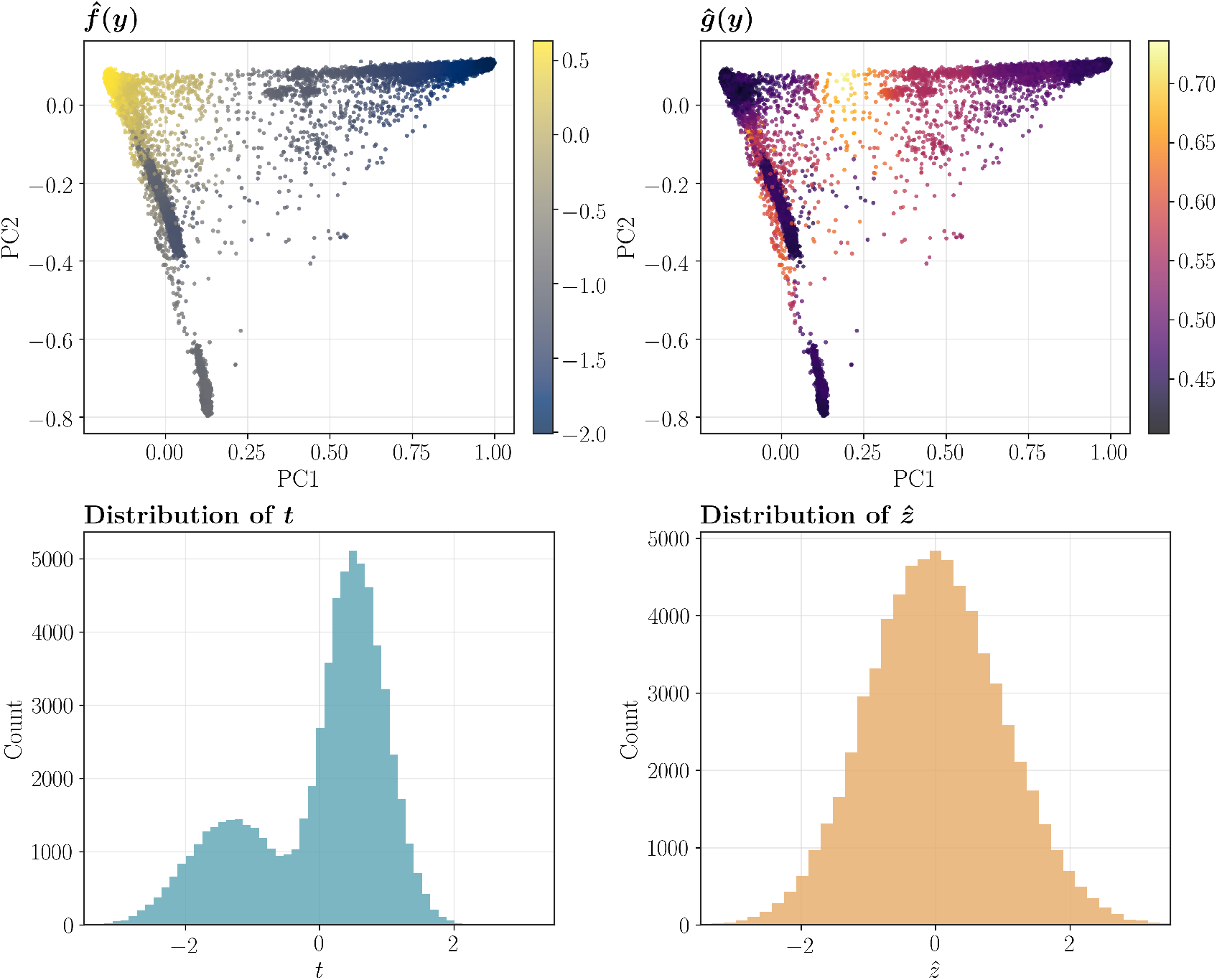
Qualitative illustration of the learned functionals. The top row shows scatter plots of the first two principal components (PC1 and PC2) of the ancestry representation, with points colored by the estimated functions 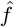(left) and 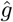 (right). These panels highlight how variation in the learned quantities is organized across the low-dimensional ancestry space. The bottom row compares the marginal distributions of t and 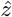: the raw score t exhibits a multimodal distribution, while the transformed variable 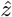 is approximately Gaussian. Together, these plots provide a qualitative view of both the dependence of 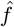 and 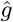 on ancestry structure and the Gaussianization effect of the learned transformation.

### Distribution of η and ρ

We evaluate the values of 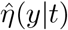 and 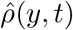 across all available PRS models. These estimates provide a measure of correlation between the raw scores *t* and the ancestry representations *y*, and in turn, an estimate of *C*. Figure 5 shows histograms of both 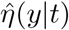 and 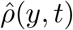 for *d* = 2 and *d* = 10, together with scatter plots comparing the two metrics. We observe that the majority of models have 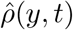 values around 0.2, although a non-negligible subset exhibits substantially larger ancestry bias, with values approaching 1. In addition, for *d* = 2, the estimates of 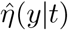 are significantly larger than 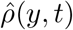, whereas for *d* = 10 the two estimates become closer. This suggests that ancestry-dependent distortions appear more linear as the dimensionality of the representation increases. Similarly, the non-linear nature of these distortions further justifies the use of more flexible non-parametric models over linear approaches.

**Figure 5.**
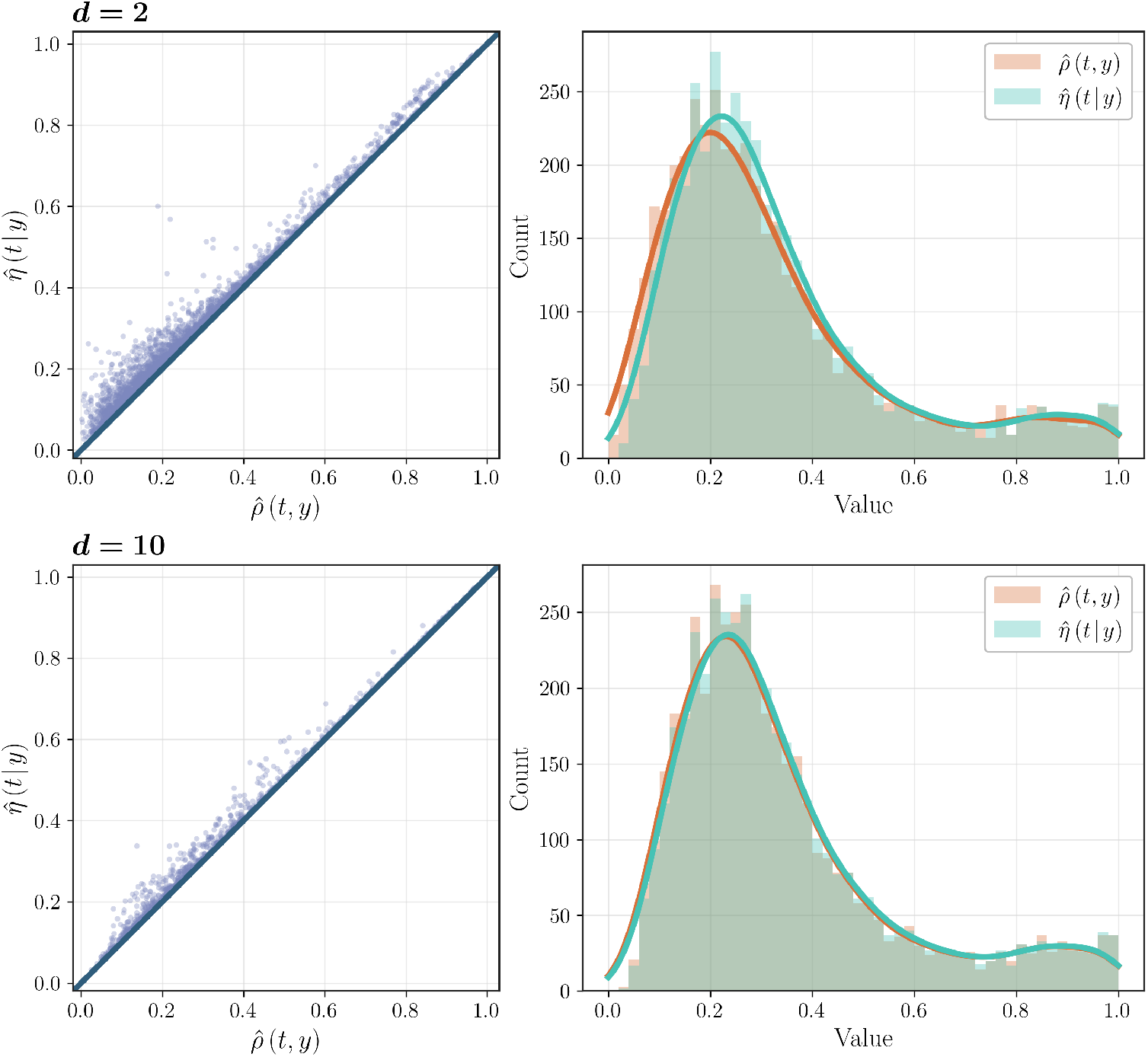
Comparison of 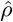 and 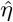 across models. The left column shows scatter plots of 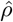 versus 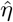, where the horizontal axis is 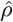 and the vertical axis is 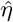. For the d = 2 setting shown in the top row, the points lie predominantly above the diagonal, indicating that 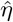 is generally larger than 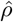. The right column shows the corresponding histograms of 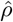 and 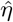, with the horizontal axis ranging from 0 to 1 and the vertical axis representing the number of models. Most models have 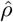 and 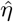 concentrated around 0.2. Both 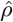 and 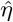 can be seen as reasonable empirical approximations of the constant C.

### Mean squared errors across K

We estimate the mean squared error lower bound MSE^LB^ over a wide range of neighborhood sizes *K*. Figure 6 shows the MSE^LB^ for all models, together with the mean and median errors. We observe a clear U-shaped behavior, consistent with theoretical predictions and simulations, with lower errors for *K* in the range of approximately 500 to 3000. The curves also exhibit step increases around *K* ≈ 3000 and *K* ≈ 10000. We hypothesize that this behavior arises as samples from different subpopulations enter the neighborhood. Specifically, for *K* ≲ 3000, most neighbors belong to the same subpopulation; for 3000 ≲ *K* ≲ 10000, admixed samples and individuals with ancestry from similar subpopulations enter the neighborhood; and for *K* ≳ 10000, the neighborhood is composed of highly diverse ancestries (e.g., African samples being adjusted using European individuals), leading to poorer adjustment.

**Figure 6.**
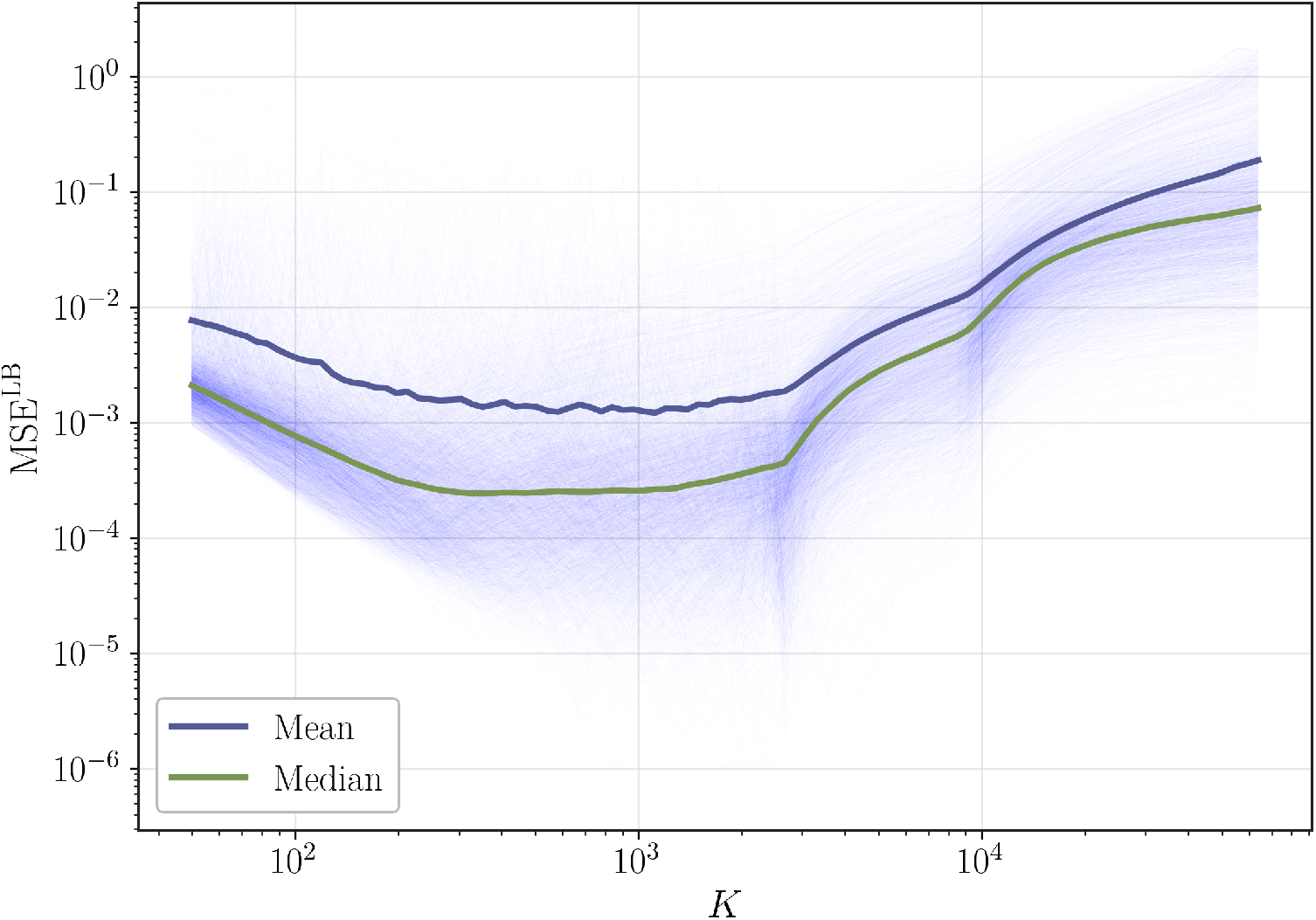
Lower-bound MSE as a function of K across models. The figure shows the empirical lower-bound mean squared error, MSE^LB^, as a function of the number of neighbors K across the considered models. As K varies, the curves exhibit several step increases, corresponding to transition points where admixed and individuals from external ancestries enter the nearest neighborhoods. Overall, the figure illustrates how MSE^LB^ follows a U-shaped curved as predicted by the theoretical bounds.

### Relationship between 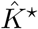 and 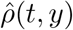

For each model, we find the optimal value of *K* that minimizes the empirical estimate of the lower bound MŜE^LB^ (Equation (4.1)) and compare it with 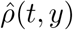, which provides an empirical lower bound of the complexity measure *C*. Figure 7 shows the relationship between 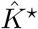 and 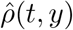 for all models in the PGSCatalog, as well as the theoretical bound derived in Corollary 3.5. Note that Corollary 3.5 is derived from the upper bound MSE^UB^, and therefore the value of *K* minimizing this bound does not necessarily coincide with the one minimizing the true MSE. Similarly, the value of *K* minimizing MSE^LB^ need not match the optimal value for the true MSE. In addition, the fact that finite-sample empirical estimates are used further distorts the estimate of the optimal *K*^⋆^. However, despite these discrepancies, the theoretical bound 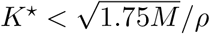 captures the expected relationship between *K*^⋆^ and *C* with surprising accuracy. Note that as *K*^⋆^ increases (or equivalently, as 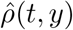 decreases), the discrepancy between the empirical estimates and the theoretical bound becomes larger. This is partly explained by the fact that the actual MSE does not change significantly between *K* values once *K*^⋆^ is significantly large.

**Figure 7.**
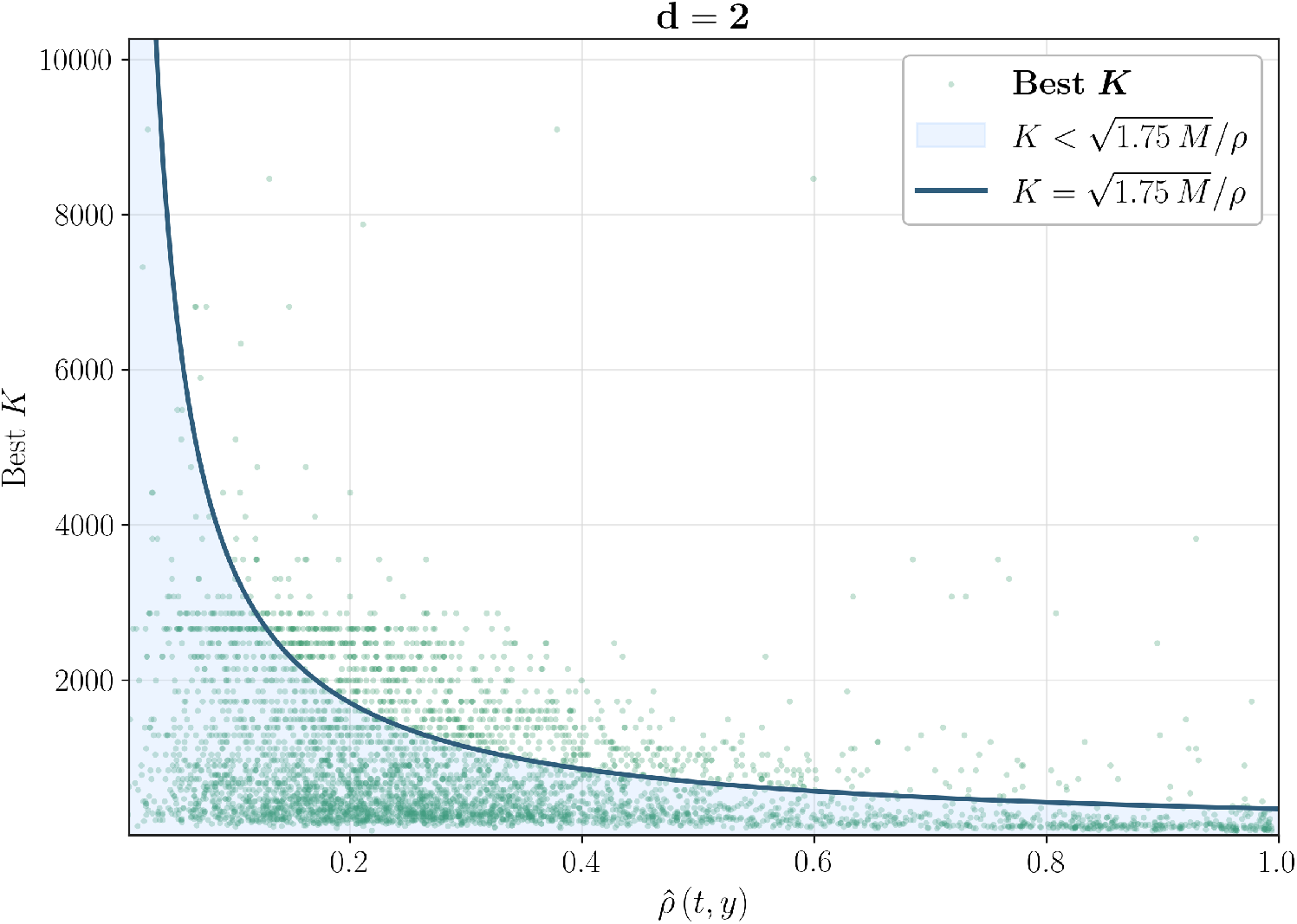
Relationship between optimal neighborhood size K^⋆^ and empirical complexity 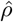. For each model in the PGSCatalog, we compute the optimal number of neighbors, 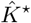, that minimizes the empirical lower-bound MŜE^LB^ from Equation (4.1), and compare it with 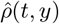, which provides an empirical lower bound of the complexity measure C. The figure also shows the theoretical relationship from Corollary 3.5, namely 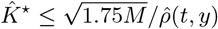.

### Benchmark with Competing Methods

We evaluate the proposed neighborhood-based (KNN) and kernel-based weighted adjustment (Kernel) methods, and compare them with competing approaches, including categorical-based adjustment (Cluster) (Section 2.2.1) [14, 10], linear adjustment (Linear) [20, 22], linear adjustment with variance parametrized as a linear mapping (Linear-Var) [18, 6, 11], and cluster-specific linear adjustment (Linear-Cl.) [3] (Section 2.2.2). For the categorical-based methods, instead of relying on predefined ethnic labels, we apply *K*-Means in the PCA space, providing a clustering that accurately captures genetic variation. To compare the performance of all methods, we use LOO estimators for each technique and perform a 50–50 split for validation and testing, where the validation set is used to select the optimal hyperparameters. The process is repeated four times, and the average metrics and standard deviations are reported. The hyperparameter settings that minimize the mean squared error lower bound MSE^LB^ on the validation set are used for evaluation on the test set. Up to 20 hyperparameter trials are performed for each method. The hyperparameter search space is chosen to reflect scenarios commonly considered in prior work. In particular, the number of clusters for the categorical-based methods ranges from 2 to 6. The linear models use a ridge regression parameter ranging from 10^−3^ to 10. For Linear-Var, we use a robust estimator for the variance term, combining clamping and ridge regularization. For the kernel method, we restrict the analysis to a Laplacian kernel with bandwidth *h* ranging from 0.005 to 0.01, and evaluate neighborhood sizes *K* ranging from 500 to 5000.

For all methods, we measure the mean squared error lower bound MSE^LB^, the empirical Pearson correlation 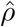, and the correlation ratio 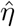, as well as the Kolmogorov–Smirnov statistic *KS* and the maximum Spearman correlation across all *d* dimensions, defined as 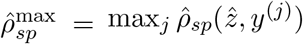, where *y*^(*j*)^ denotes the *j*th dimension of *y* and 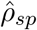 is the Spearman correlation between two real numbers. Note that for all evaluation metrics, lower values indicate better adjustment. We report results for both *d* = 2 and *d* = 10. For a given value of *d*, the same dimensionality is used for training, evaluation, and metric computation.

Table 1 reports the results (scaled by a factor of ×10^3^ for readability), with the lowest values highlighted in bold and the second lowest underlined. We observe that the Kernel method typically achieves the best adjustment, with KNN generally obtaining the second-best results across most metrics, except in a few cases where Linear-Cl ranks second. For smaller values of *d*, the gap between linear adjustment and KNN/Kernel methods is larger, indicating a stronger non-linear relationship between the ancestry representation and the distortion. This gap decreases as *d* increases. Similarly, the category-based adjustment (Cluster), which assumes a non-linear ancestry bias, outperforms linear methods when *d* = 2 but underperforms when *d* = 10. This behavior is consistent with the non-linear patterns observed in Figure 5.

**Table 1.**
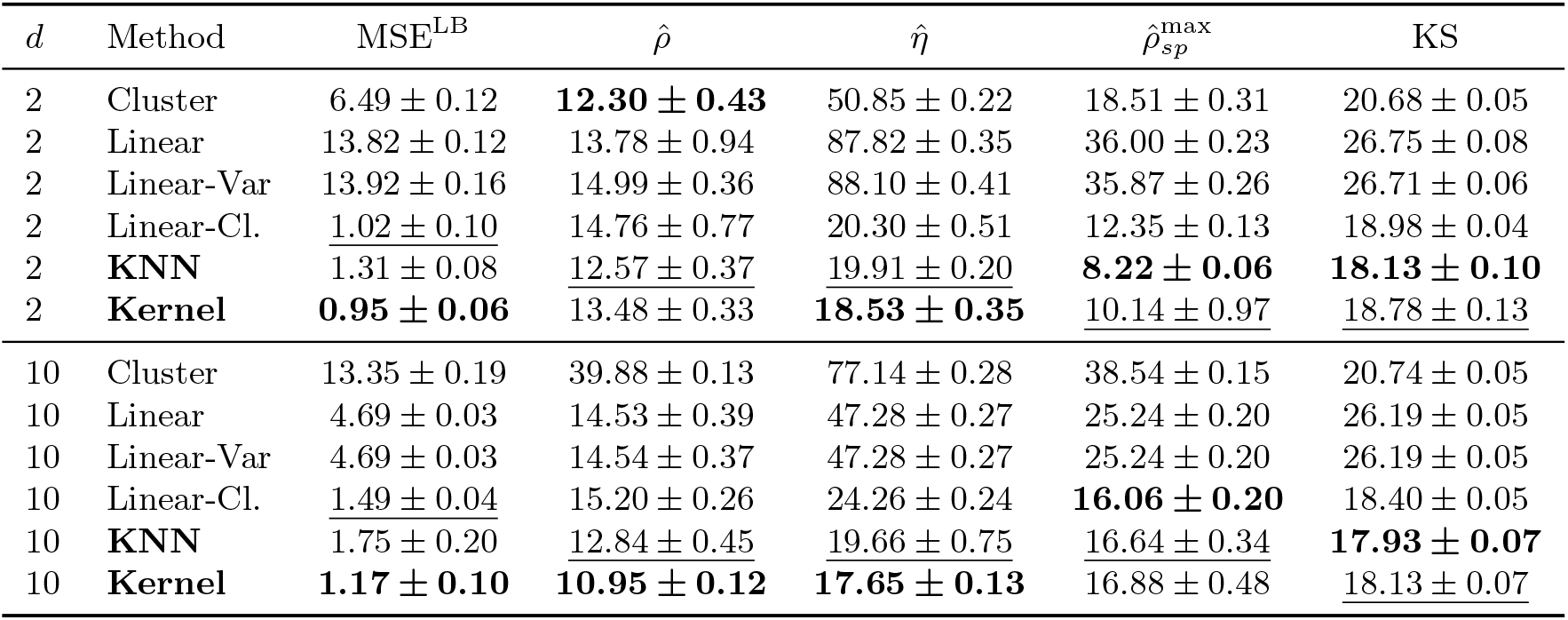
Benchmark against competing ancestry-adjustment methods. Comparison of the proposed neighborhood-based adjustment (**KNN**) and kernel-based weighted adjustment (**Kernel**) against category-based adjustment (**Cluster**), linear adjustment (**Linear**), linear adjustment with variance parametrization (**Linear-Var**), and cluster-specific linear adjustment (**Linear-Cl**.) for ancestry dimensions d = 2 and d = 10. Reported values are averaged over four validation/test splits and shown as mean ± standard deviation, scaled by a factor of ×10^3^ for readability. Hyperparameters are selected on the validation set by minimizing MSE^LB^, and all evaluation metrics follow a lower-is-better criterion. The best result in each column is highlighted in bold and the second-best is underlined.

| $d$ | Method | $\text{MSE}^{\text{LB}}$ | $\hat{\rho}$ | $\hat{\eta}$ | $\hat{\rho}_{sp}^{\text{max}}$ | KS |
| --- | --- | --- | --- | --- | --- | --- |
| 2 | Cluster | $6.49 \pm 0.12$ | **$12.30 \pm 0.43$** | $50.85 \pm 0.22$ | $18.51 \pm 0.31$ | $20.68 \pm 0.05$ |
| 2 | Linear | $13.82 \pm 0.12$ | $13.78 \pm 0.94$ | $87.82 \pm 0.35$ | $36.00 \pm 0.23$ | $26.75 \pm 0.08$ |
| 2 | Linear-Var | $13.92 \pm 0.16$ | $14.99 \pm 0.36$ | $88.10 \pm 0.41$ | $35.87 \pm 0.26$ | $26.71 \pm 0.06$ |
| 2 | Linear-Cl. | <u>$1.02 \pm 0.10$</u> | $14.76 \pm 0.77$ | $20.30 \pm 0.51$ | $12.35 \pm 0.13$ | $18.98 \pm 0.04$ |
| 2 | **KNN** | $1.31 \pm 0.08$ | <u>$12.57 \pm 0.37$</u> | <u>$19.91 \pm 0.20$</u> | **$8.22 \pm 0.06$** | **$18.13 \pm 0.10$** |
| 2 | **Kernel** | **$0.95 \pm 0.06$** | $13.48 \pm 0.33$ | **$18.53 \pm 0.35$** | <u>$10.14 \pm 0.97$</u> | <u>$18.78 \pm 0.13$</u> |
| 10 | Cluster | $13.35 \pm 0.19$ | $39.88 \pm 0.13$ | $77.14 \pm 0.28$ | $38.54 \pm 0.15$ | $20.74 \pm 0.05$ |
| 10 | Linear | $4.69 \pm 0.03$ | $14.53 \pm 0.39$ | $47.28 \pm 0.27$ | $25.24 \pm 0.20$ | $26.19 \pm 0.05$ |
| 10 | Linear-Var | $4.69 \pm 0.03$ | $14.54 \pm 0.37$ | $47.28 \pm 0.27$ | $25.24 \pm 0.20$ | $26.19 \pm 0.05$ |
| 10 | Linear-Cl. | <u>$1.49 \pm 0.04$</u> | $15.20 \pm 0.26$ | $24.26 \pm 0.24$ | **$16.06 \pm 0.20$** | $18.40 \pm 0.05$ |
| 10 | **KNN** | $1.75 \pm 0.20$ | <u>$12.84 \pm 0.45$</u> | <u>$19.66 \pm 0.75$</u> | <u>$16.64 \pm 0.34$</u> | **$17.93 \pm 0.07$** |
| 10 | **Kernel** | **$1.17 \pm 0.10$** | **$10.95 \pm 0.12$** | **$17.65 \pm 0.13$** | $16.88 \pm 0.48$ | <u>$18.13 \pm 0.07$</u> |

### Clinical Evaluation

The ancestry-adjustment of polygenic scores provides Gaussian risk estimates that are approximately independent of the genetic ancestry information and that capture the relevant polygenetic factors. Such scores are typically used as inputs to downstream risk prediction models, which also include covariates such as Principal Components, age, sex and BMI [14]. Moreover, ancestry-adjusted scores are inspected by physicians and genetic counselors, in combination with other components such as monogenic risk, family history, and socioeconomic determinants, to provide improved diagnosis and treatments. A typical application of ancestry-adjusted scores is to detect high-risk individuals by analyzing if the score falls within the high tail of the distribution (e.g. 90th or 95th percentile) which have been shown to approximately translate to a *>* 2× increased risk on European populations [11].

A common way to evaluate the clinical usefulness of the polygenic models is by measuring its predictivness for a given trait or disease in biobanks or other databases where patient-level clinical information is available. Namely, such evaluation is performed by comparing the performance of a *full model*, that includes as inputs the polygenic risk score and other covariates such as principal components, age, sex and other clinical factors, with a *base model*, where the polygenic risk score is excluded. Metrics such as the delta of the Area Under the Receiver Operating Characteristic curve (AUC-ROC), or Net Reclassification Improvement (NRI) are typically used. However, such metrics do not properly capture the effect of applying ancestry adjustment as the full model already alleviates some ancestry biases. For example, a full model based on a logistic regression that takes principal components and the risk score can implicitly learn to perform a linear ancestry adjustment. On the other hand, measuring the PRS score directly (without full and base models) with metrics such as AUC can be deceptive, as in the case where principal components are highly correlated with a given trait, a poorly adjusted model might appear to perform well, while, in fact, the model is simply correlated with the ancestry representation.

In order to provide an evaluation that highlights the importance of ancestry adjustment for clinical applications, we use two metrics that capture the quality of the score’s tail: the lift and the score exceedance error. The lift (sometimes referred to as enrichment) measures how much the prevalence of the trait is increased in the tail of the score distribution relative to its prevalence in the full population. Let *h*_*i*_ ∈ {0, 1} denote a binary label specifying whether individual *i* has the trait or disease of interest. In this evaluation, the tail is defined as the set of individuals with 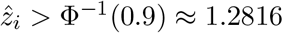, corresponding to the 90th percentile of a standard normal distribution. Therefore, the lift is expressed as

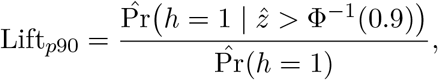

where the empirical probability is defined as 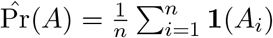, and hence

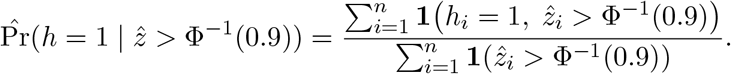

A larger value of Lift_*p*90_ indicates that the tail is able to identify high-risk individuals more accurately.

The score exceedance error measures the mismatch between the expected fraction of samples in the tail (10% for the 90th percentile tail) and the empirical fraction of samples exceeding the corresponding threshold. Namely,

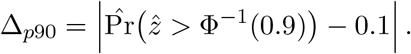

An ancestry-adjusted score that is both predictive and follows a Gaussian distribution, will lead to low Δ_*p*90_ and high Lift_*p*90_. On the other hand, large values of Δ_*p*90_ could indicate poor ancestry adjustment, leading to bi-modal distributed scores, where the tail over-represents some populations while under-representing others, making it unreliable for identifying high-risk individuals.

We evaluate all the models from the PGSCatalog for atrial fibrillation, hypothyroidism, breast cancer, colon cancer, high low density lipoprotein cholesterol, high triglycerides, and type 2 diabetes diseases. Each trait has between 10 to 80 models and we evaluate all of them performing all the described ancestry-adjustment techniques, as well as using the raw un-adjusted score. Note that for the raw score, we first perform a simple standard scaling as many scores are unit-less and their magnitudes can vary multiple orders of magnitude. The lift and exceedance errors are computed at the 90th percentile tail and we use the adjusted scores computed with the ancestry representation of dimensionality *d* = 2.

Figure 8 shows an example of the PRS models for breast cancer with metrics computed using only the Non-European individuals. We can observe a Pearson correlation between Lift_*p*90_ and Δ_*p*90_ of -0.246, indicating that non-Gaussianity and poorly calibrated tails can decrease the predictive performance of the polygenic score’s tail. In fact, by looking at all traits, adjustment and model simultaneously, we observe a Pearson correlation between Lift_*p*90_ and Δ_*p*90_ of -0.35, indicating that such behavior is shared between PRS models of different diseases and traits. We can observe that most raw scores and scores adjusted with the linear and linear-var models provide a exceedance error Δ_*p*90_ higher than 1%, with many raw scores having a Δ_*p*90_ higher than 10%. Such high exceedance errors indicate that the tails of those poorly adjusted scores can not be reliably used to identify high-risk individuals, as the tails are over- or under-representing specific ancestry groups. Namely, the distribution of the Lift_*p*90_ further characterizes such behavior where the adjustment methods of cluster, cluster-linear, and our proposed knn and kernel provide higher enrichment in the tails than the linear and unadjusted scores, while providing a lower Δ_*p*90_, with most models having a Δ_*p*90_ below 1%. Note that the highly predictive models have a Lift_*p*90_ of over 1.8, which is in line to the ×2 enrichment in tails reported in other studies [14].

**Figure 8.**
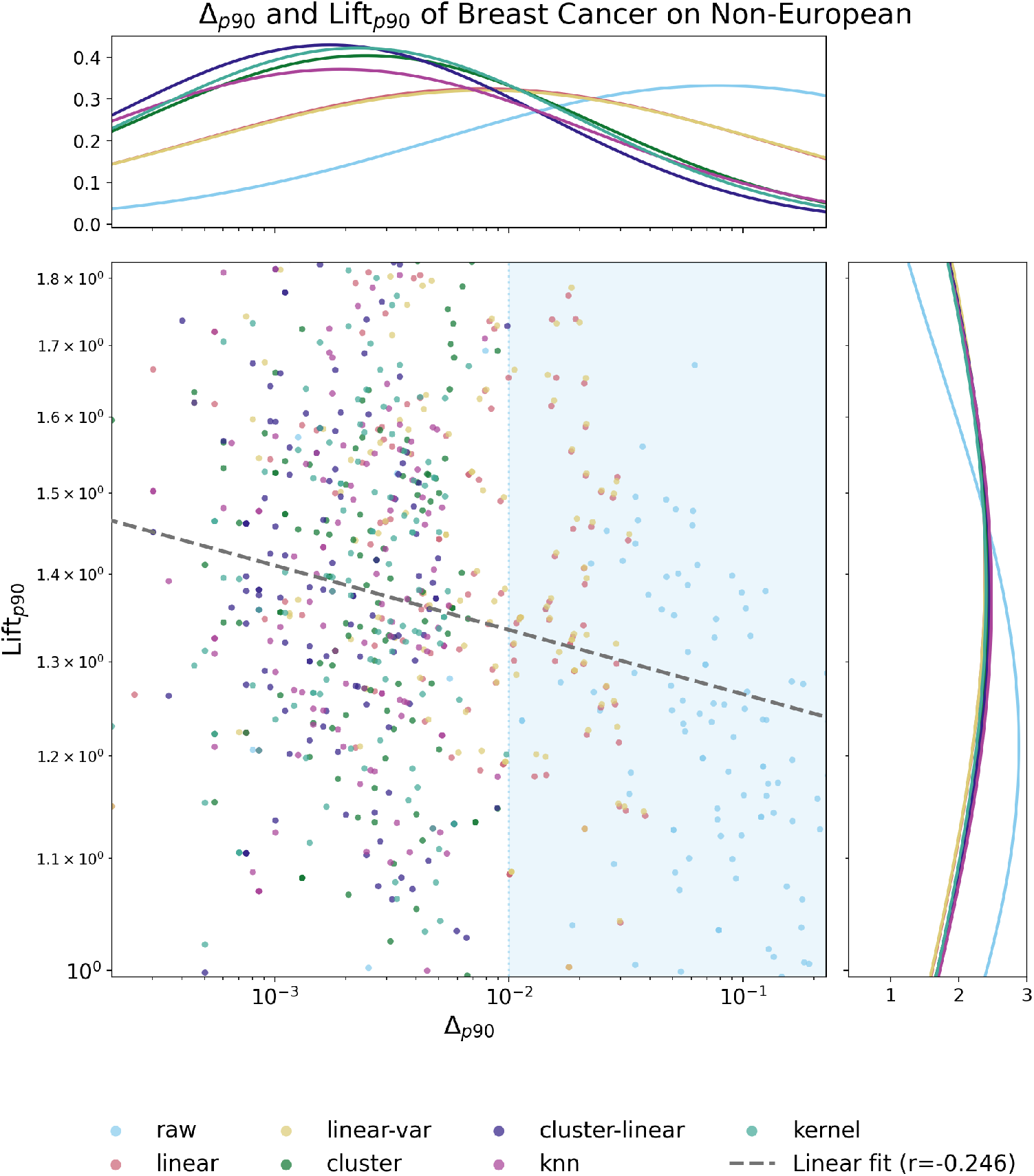
Relationship between enrichment at the tail Lift_p90_ and score exceedance error Δ_p90_ in Non-Europeans for Breast Cancer. Lift and exceedance error show a negative correlation for breast cancer models in the UK Biobank, with most of the un-adjusted and linearly-adjusted scores having a exceedance error higher than 1% (shaded area). Our proposed methods (Knn and kernel) and cluster-linear and cluster perform similarly providing both low exceedance error and higher enrichment at the 90th percentile tail.

Figure 9 shows the distribution of enrichment at the tail Lift_*p*90_ (top) and score exceedance error Δ_*p*90_ (bottom) across all reported traits for each adjustment method divided by European (left) and Non-European (right) ancestry groups. First, we can observe a very similar distribution across adjustment methods within the European ancestry group, with slightly higher lift values on the unadjusted scores. The median lift across all adjustment scores is close to 2, with many models having a lift higher than 3, similar to previously reported enrichment levels [14]. Within Non-Europeans, the unadjusted (raw) score performs on average significantly worse than the adjusted scores, with a median lift significantly lower than the adjusted score’s lift. Importantly, the median lift on the Non-European group is lower than the lift of the European group across all adjustment methods, reflecting the lack of transferability of many polygenic models across population groups and highlights the need of the inclusion of more diverse populations in genomic studies. The distributions of Δ_*p*90_ show that the unadjusted scores have very large exceedance errors followed by the linear and linear-var adjustments, indicating that such tails are poorly calibrated. Note that some unadjusted models have higher lift values than their adjusted versions. Such effect can be explained by the fact that some models are highly correlated with the ancestry representation, and due to dataset-specific and socioeconomic biases, the ancestry representation can be correlated with a given trait. Namely, poorly adjusted models (or unadjusted models), which are highly correlated with ancestry, might show a large enrichment on the tails while in fact not truly capturing polygenic risk factors. Therefore, to properly characterize the capability of an adjusted score to capture the underlying genetic factors, it is important to simultaneously evaluate its independence from ancestry representations and its predictive power.

**Figure 9.**
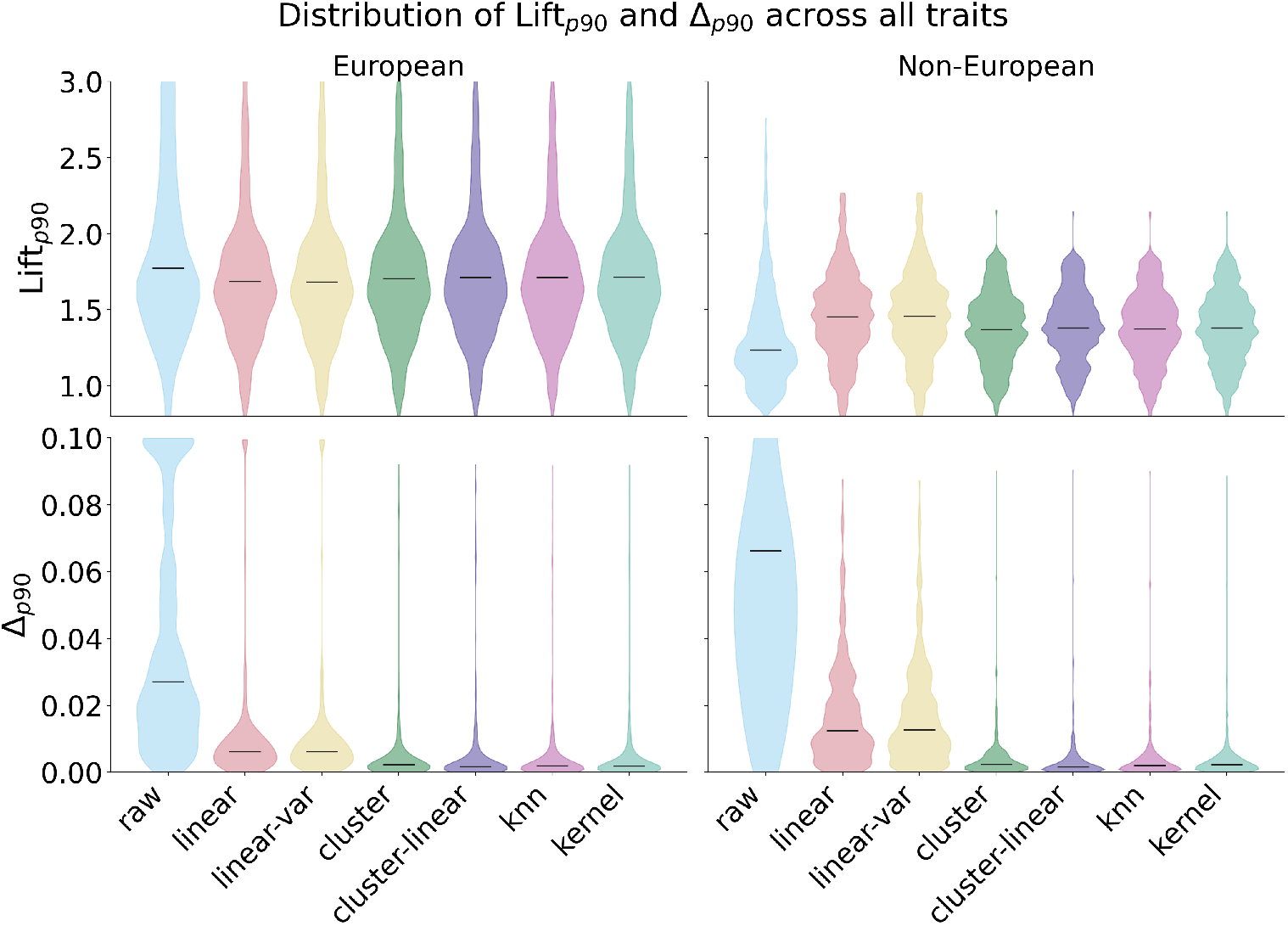
Distribution of enrichment at the tail Lift_p90_ (top) and score exceedance error Δ_p90_ (bottom) across traits for each adjustment method for Europeans (left) and Non-Europeans (right). The unadjusted score (raw), linear, and linear-var show a large Δ_p90_ in both European and Non-European The adjustment methods of cluster, cluster-linear, Knn, and kernel provide both low Δ_p90_ and high Lift_p90_. The Lift_p90_ of Non-Europeans is typically lower than the Lift_p90_ of European groups.

Figure 10 shows the average Lift_*p*90_ and Δ_*p*90_ as well as the distribution of Lift_*p*90_ for each trait in Non-European individuals. The average Lift_*p*90_ of the raw score is significantly lower than the average Lift_*p*90_ of the adjusted scores. Similarly the average Δ_*p*90_ of the raw score is significantly larger than the average Δ_*p*90_ of the adjusted scores, highlighting the importance of ancestry adjustment in order to obtain interpretable and predictive tails of the distribution. The adjustment methods of linear and linear-var provide an interesting behavior: they provide a higher average Lift_*p*90_ than cluster, cluster-linear, knn, and kernel, while providing a significantly larger average Δ_*p*90_. Therefore, the higher average Lift_*p*90_ of linear and linear-var is likely due to residual ancestry information that is correlated with the trait information leading to both increased enrichment but large score exceedance error. In terms of average Δ_*p*90_ and average Lift_*p*90_, the adjustment methods cluster, cluster-linear, Knn, and kernel, perform similarly, with the cluster method slightly under-performing. The kernel adjustment provides the largest average Lift_*p*90_. Figure 10 (bottom) shows the distribution for each trait for the cluster-linear, Knn, and kernel methods, showing similar performance. For multiple traits, the distribution of Lift_*p*90_ is highly multi-modal, with groups of models at Lift_*p*90_ values close to 1 while groups of models tending to Lift_*p*90_ values close to 1.8. Such multi-modality reflects the heterogeneity of models present in the PGSCatalog, with some models focusing mainly on European populations and under-performing in Non-European cohorts, while other methods providing competitive enrichment in Non-European cohorts.

**Figure 10.**
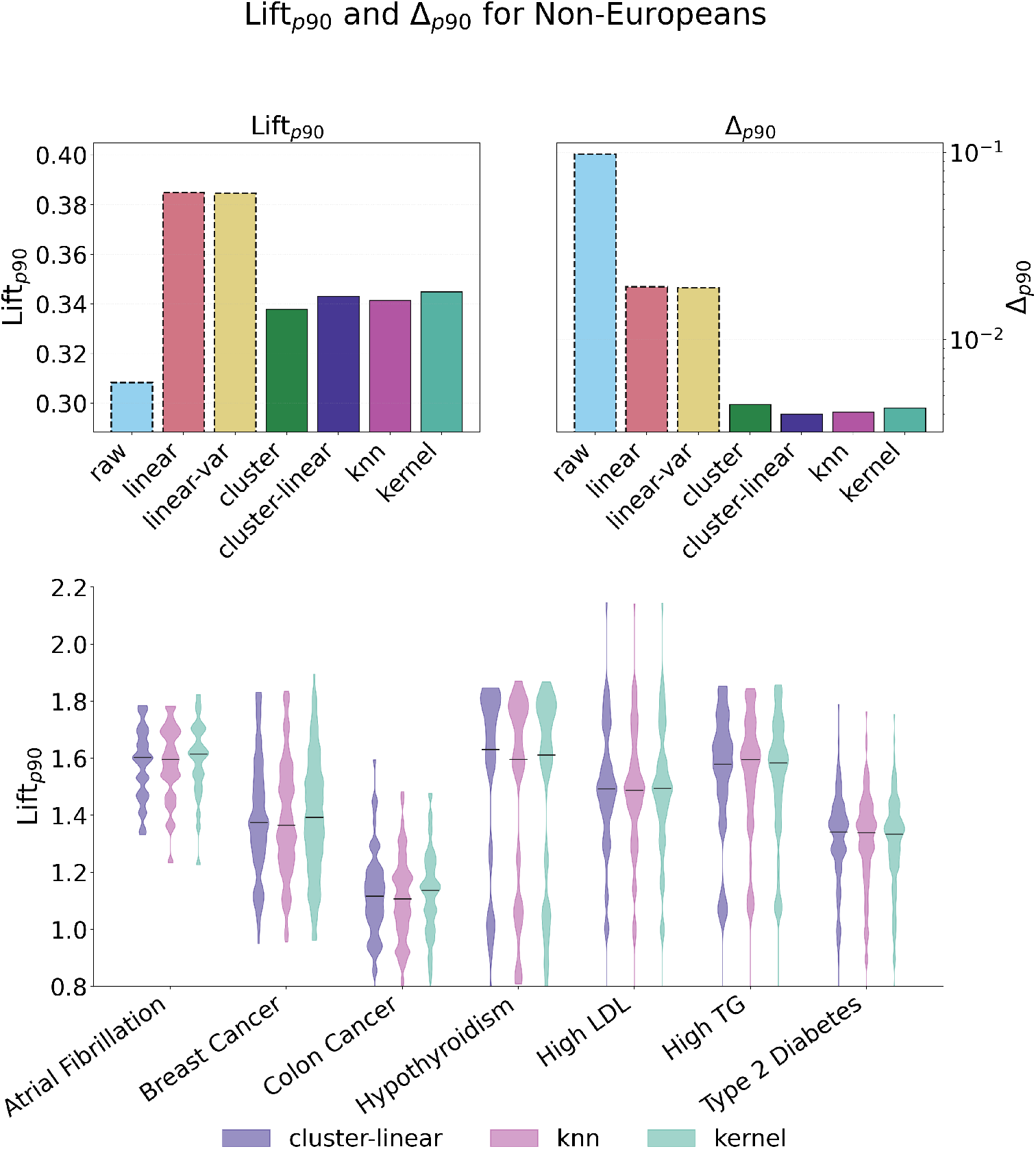
(top) Average Lift_p90_ and Δ_p90_ in Non-Europeans and (bottom) per-trait distribution of Lift_p90_. Unadjusted scores (raw) provide low Lift_p90_ and high Δ_p90_, while cluster-linear, Knn, and kernel provide both high Lift_p90_ and small Δ_p90_.

## 7. Discussion

The proposed theoretical framework highlights that ancestry adjustment for polygenic risk scores is most naturally formulated as a conditional standardization problem over a continuous ancestry representation. Under the location-scale model *t* = *z g*(*y*) + *f* (*y*), the ideal adjustment is the unique affine transformation that removes the ancestry-dependent mean and variance. This perspective places several existing methods within a common framework: category-based adjustment corresponds to a piecewise-constant estimator of *f* and *g*, linear residualization corresponds to a parametric estimator, and the proposed neighborhood and kernel methods correspond to non-parametric local estimators. The advantage of the latter is precisely that they adapt to smooth but nonlinear ancestry effects, which frequently arise in practice, especially for admixed individuals and along continuous genetic ancestry gradients.

Our empirical results are consistent with this interpretation. In the UK Biobank experiments, raw PRS often retains substantial dependence on ancestry even after linear or category-based correction, whereas neighborhood-based methods more effectively whiten the score distribution across genetic ancestry space. This behavior is also reflected in the estimated dependence measures: for many models, the empirical correlation ratio exceeds the Pearson correlation, indicating that a substantial component of the ancestry effect is non-linear. From this perspective, the proposed non-parametric estimators are not merely more flexible predictors, but methods better aligned with the geometry of the underlying nuisance structure.

An important limitation, however, is that the theory is expressed in terms of an ancestry descriptor *y*, whereas in practice only a chosen representation of ancestry is available. The variable that truly governs the distortion of the raw score may be a latent quantity *y*^⋆^, while the adjustment is performed using another representation 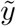. These two variables need not coincide, nor share the same dimensionality. This mismatch is conceptually important: if 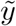 is lower-dimensional than the relevant latent ancestry variable, then some ancestry-dependent components of *f* and *g* may be unobserved. In such cases, even a statistically optimal estimator with respect to 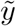 may leave residual ancestry bias uncorrected, simply because the adjustment variable does not contain sufficient information. Furthermore, the quantities *ρ, η*, Spearman correlation, and the empirical MSE bounds are all defined relative to the chosen ancestry representation. Consequently, small empirical dependence with respect to 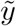 does not imply that the adjusted score is truly ancestry-independent in a broader sense; it only indicates weak dependence on the specific representation being tested. It is therefore possible that no detectable correlation remains with the chosen low-dimensional predictor, while a richer or more appropriate ancestry representation would still reveal substantial residual dependence. In this sense, empirical validation of ancestry adjustment is inseparable from the choice of ancestry coordinates used both for correction and evaluation.

The dimensionality *d* of the ancestry representation also directly affects the statistical behavior of the proposed non-parametric estimators. The bounds exhibit the usual curse of dimensionality through the bias term (*K/M*)^2*/d*^, implying that larger *d* requires substantially more reference samples to achieve the same local resolution. Thus, increasing *d* may improve representational fidelity but worsens sample complexity. This creates a practical trade-off: low-dimensional representations may miss relevant ancestry structure, whereas high-dimensional representations may render local estimation unstable unless very large and diverse reference panels are available.

There is also a second failure mode when the ancestry representation is overly expressive. In the extreme, one could use the full genotype *g*, or a representation rich enough to capture not only ancestry effects but also the biological signal that the PRS is intended to measure. In this regime, the adjustment procedure may remove not only nuisance variation due to ancestry, but also genuine risk-related variation, resulting in an over-corrected and biologically distorted score. Namely, if the adjustment covariate contains the target signal, the decomposition between nuisance structure and true score is no longer identifiable. This suggests that ancestry adjustment should rely on representations that capture population structure while remaining sufficiently disentangled from the disease-specific signal of interest.

These limitations are not unique to the proposed methods but are applicable to the ancestry adjustment problem as whole. Ancestry adjustment is ultimately performed to decouple ancestry-dependent effects, which might capture undesired socioeconomic biases, dataset imbalances, or algorithmic artifacts, from true polygenic effects that can be interpreted by physicians and genetic counselors, among others, in order to improve diagnosis, detect high risk individuals, and provide personalized treatments. However, the boundaries dividing “ancestry” and “polygenic” factors are not well defined, and a strict clear division might be in fact impossible. Therefore, a “true” optimal ancestry representation is not just inaccessible, but likely inexistent. Therefore, when performing ancestry-adjusted PRS, practitioners should understand the limitations of ancestry adjustment and how the nature of the ancestry representations affect the final polygenic risk score.

## 8. Conclusions

We developed a unified theoretical framework for ancestry adjustment in polygenic risk scores by modeling the raw score as a Gaussian location–scale transformation whose mean and dispersion depend on a continuous ancestry descriptor. Within this framework, the ideal adjustment is the unique affine standardization that removes ancestry-dependent mean and variance effects, reducing the problem to the estimation of conditional moments. This perspective clarifies why widely used approaches—category-based standardization and linear residualization—can be effective under their respective structural assumptions, yet may leave substantial residual ancestry dependence when ancestry varies continuously or individuals are admixed. To address these limitations, we introduced neighborhood-based non-parametric adjustment methods that estimate local mean and scale in ancestry space via *K*-nearest neighbors and kernel-weighted variants. We derived finite-sample error bounds that decompose bias and variance as functions of *K* and the reference set size, yielding consistency and an explicit choice of *K* that optimizes the theoretical upper bound under mild smoothness assumptions. Empirically, experiments on large biobank-scale resources demonstrate that neighborhood-based adjustment more effectively removes ancestry-associated shifts than category-based or linear methods, resulting in better-calibrated standardized scores across ancestry gradients and supporting more equitable interpretation of PRS in diverse and admixed populations.

## Data Availability

All data presented in the work can be generated from the UK Biobank.

## Appendix A Asymptotic Analysis of Traditional Ancestry-adjustment Methods

### A.1. Affine Models: Optimal Conditional Standardization

#### Proposition A.1

(Optimal affine adjustment). *Under the generative model* (2.1) *with g*(*y*) *>* 0, *consider affine adjustments of the form* (2.2). *If* 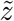 *satisfies* 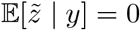 *and* 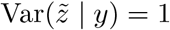 *for all y, then necessarily*

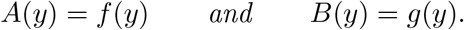

*In particular, the unique affine standardization achieving conditional centering and unit variance is* 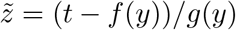.

*Proof*. From (2.1) we have E[*t* | *y*] = *f* (*y*) and Var(*t* | *y*) = *g*^2^(*y*). Plugging into (2.2), the constraints 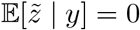 and 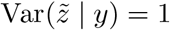 imply 0 = (*f* (*y*)−*A*(*y*))*/B*(*y*) and 1 = *g*^2^(*y*)*/B*^2^(*y*). Since *B*(*y*) *>* 0, it follows that *A*(*y*) = *f* (*y*) and *B*(*y*) = *g*(*y*).

### A.2. Categorical ancestry adjustment: asymptotic optimality

Throughout, 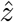 is evaluated on a fresh draw (*t, y*) independent of the data used to form 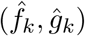 in (B.5)–(2.4).

#### Lemma A.2 (Consistency of within-category estimators).

*Fix a category k with* ℙ(*C*(*y*) = *k*) *>* 0. *Under the categorical model of Section 2.2.1, as n*_*k*_ → ∞,

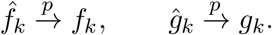

*Proof*. Within category *k*, {*t*_*i*_ : *i* ∈ *I*_*k*_} are i.i.d. with finite second moment. Hence 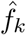 is consistent by the law of large numbers. The unbiased sample variance 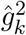 is consistent for 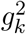 by standard variance consistency, and continuity of the square root yields *ĝ*_*k*_ → *g*_*k*_ in probability.

#### Lemma A.3

(*L*^2^ convergence within a fixed category). *Fix a category k and consider the out-of-sample score* 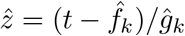 *and the ideal score z* = (*t* − *f*_*k*_)*/g*_*k*_. *If n*_*k*_ → ∞, *then*

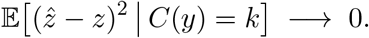

*Proof*. Write

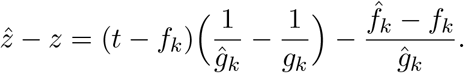

Condition on 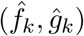 and use out-of-sample independence so that *t* is independent of 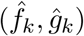 given *C*(*y*) = *k*. Since E[*t* − *f*_*k*_ | *C*(*y*) = *k*] = 0, the cross term vanishes and

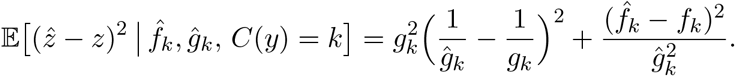

By Lemma A.2, 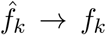 and *ĝ*_*k*_ → *g*_*k*_ in probability; since *g*_*k*_ *>* 0, also 1*/ĝ*_*k*_ → 1*/g*_*k*_ in probability. Under the categorical model (Gaussian within each category), these terms are uniformly integrable; hence, their expectations converge to 0, yielding the claim.

#### Proposition A.4 (Asymptotic optimality across categories)

*Assume that for every category k with* ℙ(*C*(*y*) = *k*) *>* 0 *we have n*_*k*_ → ∞. *Then*

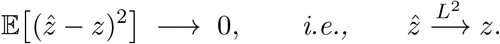

*Proof*. By the law of total expectation and Lemma A.3,

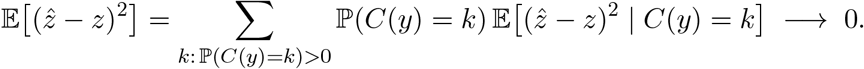

#### Corollary A.5 (Asymptotic ancestry removal)

*Under the assumptions of Proposition A.4, for any k with* ℙ(*C*(*y*) = *k*) *>* 0,

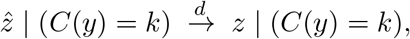

*and the limiting distribution is the same for all such k*.

*Proof*. Lemma A.3 implies 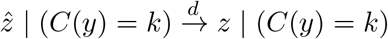. Under the categorical model in the main text, the conditional distribution of *z* is identical across categories by construction; hence, the limit does not depend on *k*.

### A.3. Linear ancestry adjustment: asymptotic optimality under constant scale

We prove that under the correctly specified constant-scale linear model (2.6), the plug-in adjusted score (obtained by combining (2.7) and (2.8) in the standardization (2.2)) is asymptotically equivalent to the ideal score *z*. Throughout, 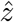 is evaluated on a fresh draw (*t, y*) independent of the sample used to fit 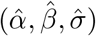.

#### Lemma A.6 (Consistency of OLS mean and variance estimators)

*Assume* (2.6). *Suppose* E∥*y*∥^2^ *<* ∞ *and* E[*yy*^⊤^] *is nonsingular, and that n* → ∞ *with p fixed. Then*

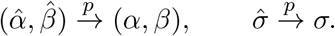

*Proof*. These are standard OLS consistency results under correct specification and finite second moments; see, e.g., any asymptotic theory for linear regression with random design. Consistency of 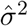 follows from consistency of the residual sum of squares per degree of freedom under the same conditions, and 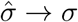 by continuity.

#### Lemma A.7 (Conditional convergence of the plug-in standardization).

*Let* 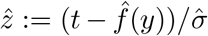 *with* 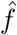 *defined in* (2.7) *and* 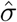 *in* (2.8). *Under the conditions of Lemma A.6 and out-of-sample evaluation, for every fixed y*,

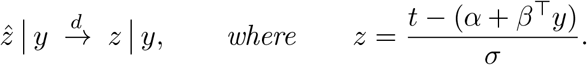

*Proof*. Write

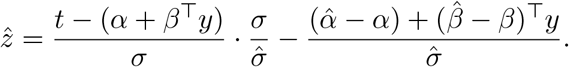

Under out-of-sample evaluation, *t* is independent of 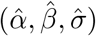 conditional on *y*. By Lemma A.6, 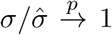 and the second term converges to 0 in probability for fixed *y*. Slutsky’s theorem yields the claim.

#### Proposition A.8 (Asymptotic optimality)

*Under the conditions of Lemma A.6 and out-of-sample evaluation, the plug-in adjusted score* 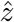 *converges to the ideal score z in distribution:*

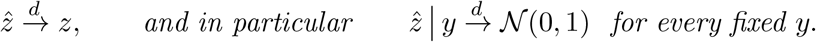

*Proof*. The first statement follows by mixing the conditional convergence in Lemma A.7 over the law of *y*. The second statement follows from (2.6), which implies *z* | *y* ∼ *N* (0, 1) for every *y*.

#### Corollary A.9 Asymptotic ancestry removal

*Under the conditions of Proposition A.8, the limiting law of* 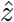 *does not depend on ancestry: for every fixed* 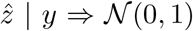, *hence the limit is identical across all ancestries*.

*Proof*. This is immediate from Proposition A.8.

## Appendix B Moment bounding of 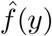 and *ĝ*(*y*).

### B.1. Basic Definitions

*Set-up and notation*. Fix an integer *K* ≥ 2. For each neighbor *i* = 1, …, *K*, we observe a pair (*z*_*i*_, *x*_*i*_) where 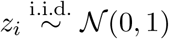, and *x*_*i*_ is arbitrary (fixed or random). Fix a reference point *y* and define the local deviations

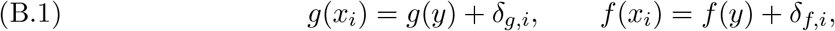

where *g*(*y*) *>* 0. The neighbor observation is

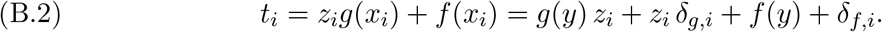

Throughout, *g*(*y*) and *f* (*y*) are treated as fixed constants. We assume block independence 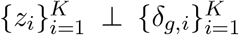 and 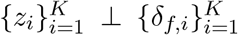, while allowing arbitrary dependence across *i* within each *δ*-block. For reference, at the target point *y* we also consider the “clean” draw

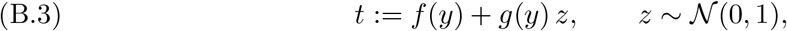

with *z* independent of the neighbor block when joint statements are needed.

*Averages and local estimators*. Define the empirical average of the Gaussian terms

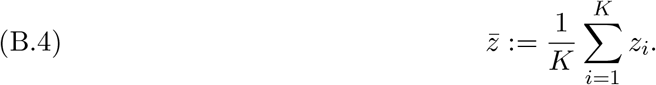

Define the local shift estimator (empirical mean of the neighbor responses)

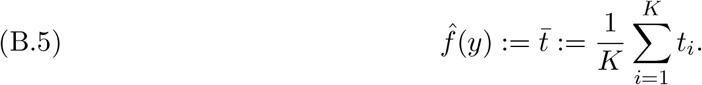

Let the empirical variance and standard deviation of 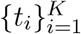 be

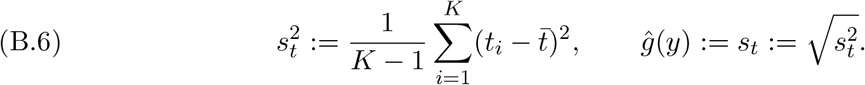

For stabilization of normalized quantities, define the lower-bounded scale *s*_*ε*_ as

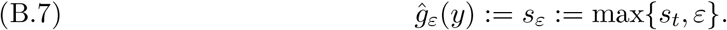

Here and throughout, *ε >* 0 denotes a fixed stabilization level.

*Moment assumptions on local deviations*. Fix nonnegative Lipschitz constants *L*_*f*_, *L*_*g*_, a scale *σ >* 0, and let *p* ∈ {1, 2, 4}. Assume averaged *p*th-moment control:

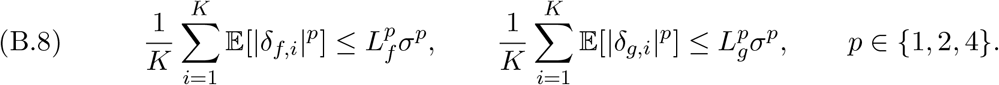

These conditions quantify typical local variation of *f* and *g* within the *K*-neighbor neighborhood, without imposing independence across neighbors.

#### B.2. Upper bounding 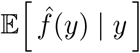

##### Lemma B.1 (Conditional mean of the local shift estimator)

*Under the set-up above*,

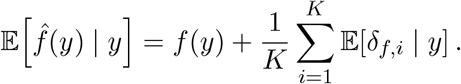

*Proof*. By linearity and the decomposition of *t*_*i*_,

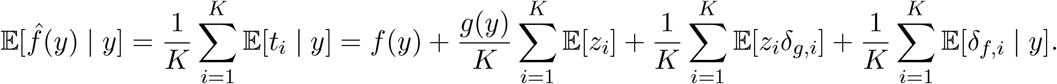

Since E[*z*_*i*_] = 0 and {*z*_*i*_} ⊥ {*δ*_*g,i*_}, we have E[*z*_*i*_*δ*_*g,i*_] = 0, which yields the claim.

##### Proposition B.2 (Mean deviation bound)

*Assume the p* = 1 *moment condition in* (B.8). *Then*

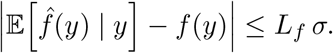

*Proof*. By Lemma B.1 and triangle inequality,

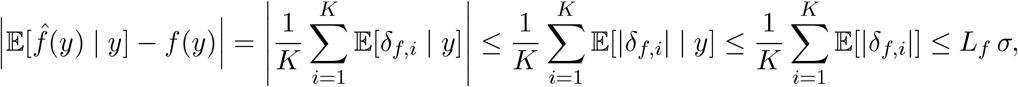

where the last step uses (B.8) with *p* = 1.

##### Corollary B.3 (Squared bias and unbiasedness)

*Under the p* = 1 *moment condition in* (B.8),

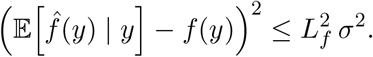

*Moreover, if* E[*δ*_*f,i*_ | *y*] = 0 *for all i, then* 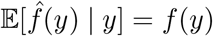.

#### B.3. Upper bounding 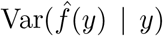

We begin by isolating the random fluctuation of 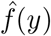 around its conditional mean.

##### Lemma B.4 (Variance decomposition)

*Under the set-up of §*(B.2)*–*(B.8),

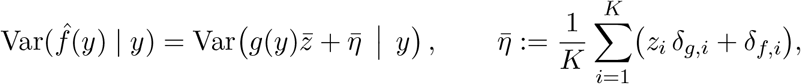

*and consequently*

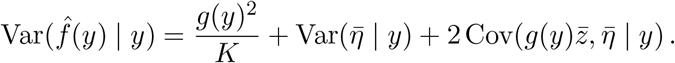

*Proof*. From (B.2) and (B.5),

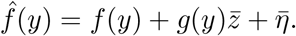

Since *f* (*y*) is deterministic, it does not contribute to the variance, giving the first identity. Expanding the variance of a sum yields the second display, and 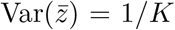 follows from 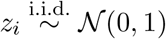.

##### Lemma B.5 (Second moment bound for 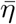).

*Assume the p* = 2 *moment condition in* (B.8). *Then*

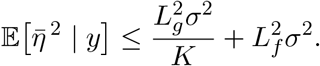

*In particular*, 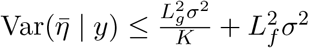.

*Proof*. Write 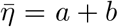 with 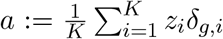 and 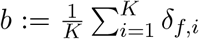. By block independence and E[*z*_*i*_] = 0, E[*ab* | *y*] = 0, hence 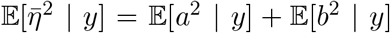. Moreover, using E[*z*_*i*_*z*_*j*_] = **1**{*i* = *j*},

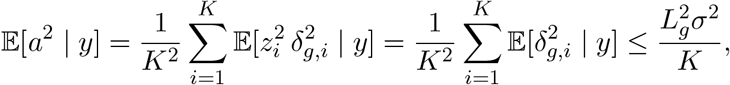

where the last step uses (B.8) with *p* = 2. For *b*, without assuming any dependence across *i*, Cauchy-Schwarz gives 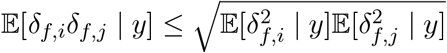, hence

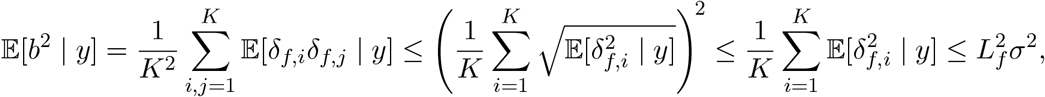

where the middle inequality is Jensen for the concave map 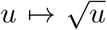, and the last step uses (B.8) with *p* = 2. The variance bound follows from 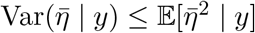.

##### Lemma B.6 (Mixed covariance bound)

*Assume the p* = 2 *moment condition in* (B.8). *Then*

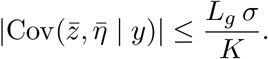

*Proof*. Since 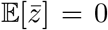, we have 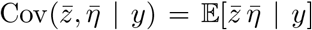. Expanding 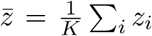 and 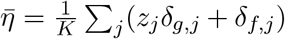, block independence and 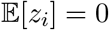 imply that all terms vanish except those with *i* = *j* and the *z*_*j*_*δ*_*g,j*_ component, yielding

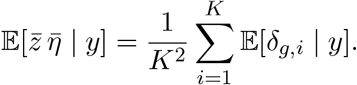

By Cauchy–Schwarz, 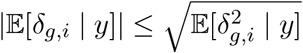, hence

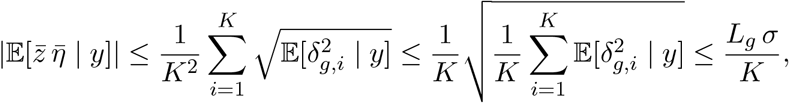

where the middle inequality is Jensen and the last step uses (B.8) with *p* = 2.

##### Proposition B.7

(Variance bound for 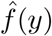. *Assume the p* = 2 *moment condition in* (B.8). *Then*

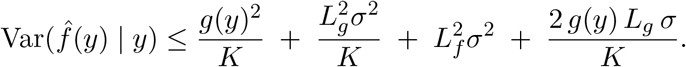

*Proof*. Combine Lemma B.4 with Lemma B.5 and Lemma B.6, using 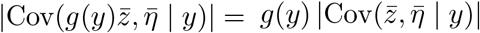 and 2Cov(·) ≤ 2|Cov(·)|.

#### B.4. Lower bounding 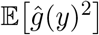

Recall *ĝ*(*y*) = *s*_*t*_ from (B.6). Throughout this subsection we assume *g*(*y*) *>* 0 and write (*x*)_+_ := max{*x*, 0}.

*Centered residual decomposition*. By (B.2) and (B.5), the centered residuals admit 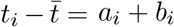, where we define the empirical means

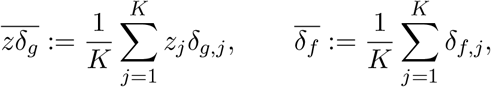

and set

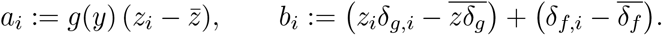

##### Lemma B.8 (Master identity for 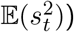.

*With* 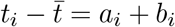 *as above*,

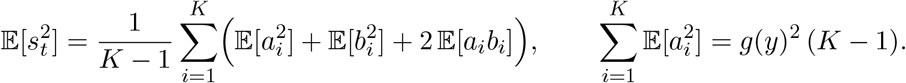

*Proof*. Expand 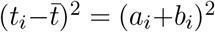 inside the definition of 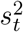 in (B.6) and take expectations. The identity for 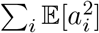 follows from 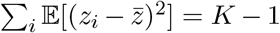 for i.i.d. standard Gaussians.

##### Proposition B.9 (Lower bound on E(*ĝ*(*y*)^2^))

*Assume the p* = 1 *moment condition in* (B.8) *for* 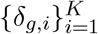. *Then*

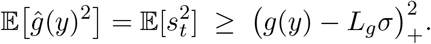

*Proof*. Using Lemma B.8, dropping the nonnegative contribution coming from *δ*_*f*_, and evaluating the Gaussian terms via the block-independence in §(B.2)–(B.8), and using E[*δ*_*g,i*_] ≥ −E[|*δ*_*g,i*_|], yields the lower-side reduction

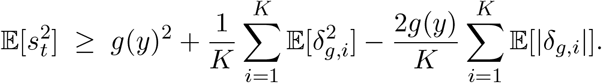

Finally, Cauchy-Schwarz 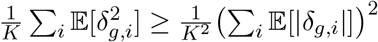, hence

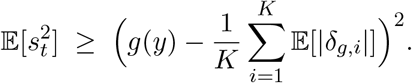

Applying (B.8) with *p* = 1 yields 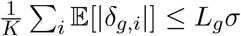, which yields the claim.

##### Proposition B.10 (Factorized form).

*Under the assumptions of Proposition B.9 and g*(*y*) *>* 0,

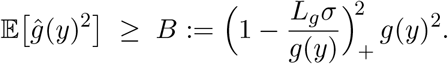

##### Corollary B.11 (Dimensionless lower bound)

*Under the assumptions of Corollary B.10 and g*(*y*) *>* 0, *let L* := max{*L*_*f*_, *L*_*g*_} *and define* 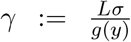. *Then the factorized lower bound B from Corollary B.10 satisfies*

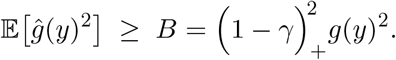

#### B.5. Upper bounding E [*ĝ*(*y*)^2^ ]

Recall *ĝ*(*y*) = *s*_*t*_ from (B.6). Reuse the centered residual decomposition from the previous subsection, namely 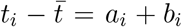 with 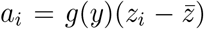 and *b*_*i*_ = *b*_*g,i*_ + *b*_*f,i*_, where 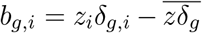 and 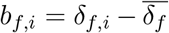. By Lemma B.8, it remains to upper bound the deviation and cross contributions.

##### Lemma B.12 (Deviation contribution).

*Under the block-independence assumptions in §*(B.2)*–* (B.8),

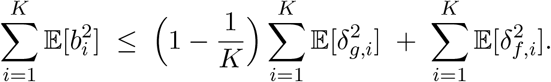

*Proof*. Since *b*_*i*_ = *b*_*g,i*_ + *b*_*f,i*_, block independence and E[*z*_*i*_] = 0 imply E[*b*_*g,i*_*b*_*f,i*_] = 0, hence 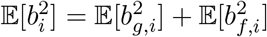. For the *g*-part, with *u*_*i*_ := *z*_*i*_*δ*_*g,i*_ and 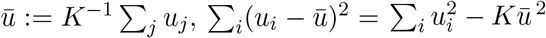, and taking expectations yields 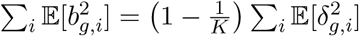. For the *f* -part, 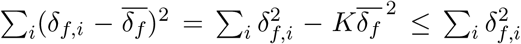 deterministically; taking expectations gives the stated bound.

##### Lemma B.13 (Cross contribution)

*Under the same assumptions*,

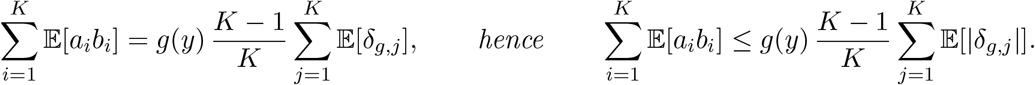

*Proof*. Decompose *b*_*i*_ = *b*_*g,i*_ + *b*_*f,i*_. The *f* -part vanishes after summation because 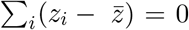 deterministically. For the *g*-part, expand 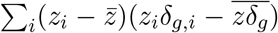 and use 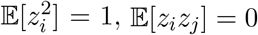 for *i* ≠ *j*, together with block independence, to obtain the identity. The inequality follows from E[*δ*_*g,j*_] ≤ E[|*δ*_*g,j*_|] and *g*(*y*) *>* 0.

##### Proposition B.14 (Raw upper bound for E(*ĝ*(*y*)^2^))

*Under the assumptions of Lemmas B.12 – B.13*,

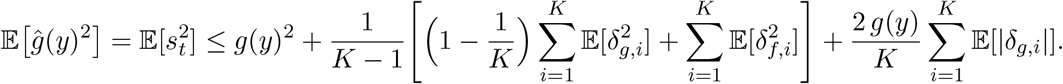

*Proof*. Start from Lemma B.8. Apply Lemma B.12 to control 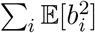 and Lemma B.13 to control ∑_i_ E[*a*_*i*_*b*_*i*_]. Using 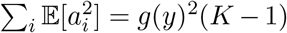 from Lemma B.8 and dividing by *K* – 1 yields the claim.

##### Proposition B.15

(Moment upper bound). *Assume* (B.8) *with p* = 1 *for* {*δ*_*g,i*_} *and p* = 2 *for* {*δ*_*g,i*_}, {*δ*_*f,i*_}. *Then, for all K* ≥ 2,

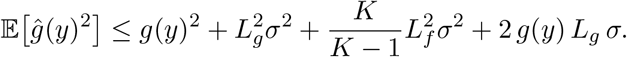

*Proof*. Apply Proposition B.14 and bound each term using (B.8):

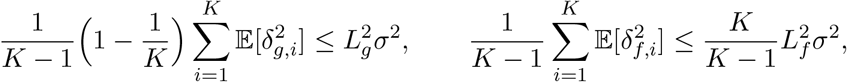

and 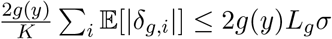. Combining yields the result.

Corollary B.16 (Simplified bound for *K* ≥ 10). *Under the assumptions of Proposition B.15, if K* ≥ 10 *then*

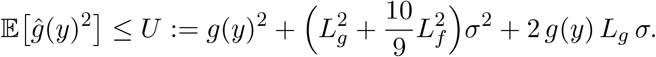

##### Corollary B.17 (Dimensionless upper bound on excess second moment)

*Under the assumptions of Corollary B.16, let L* := max{*L*_*f*_, *L*_*g*_} *and define* 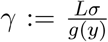 *(with g*(*y*) *>* 0*). Then, for K* ≥ 10,

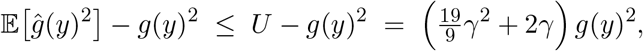

#### B.6. Upper bounding Var(*ĝ*(*y*)^2^)

Recall 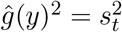 from (B.6). Write

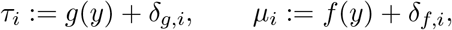

so that *t*_*i*_ = *τ*_*i*_*z*_*i*_ + *µ*_*i*_ by (B.2). Throughout we use the block-independence from §(B.2)–(B.8) and the moment controls (B.8) with *p* ∈ {1, 2, 4 }.

*Quadratic-form representation*. Let *z* := (*z*_1_, …, *z*_*K*_)^⊤^, *τ* := (*τ*_1_, …, *τ*_*K*_)^⊤^, *µ* := (*µ*_1_, …, *µ*_*K*_)^⊤^, **1** be the all-ones vector, and 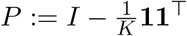. Define

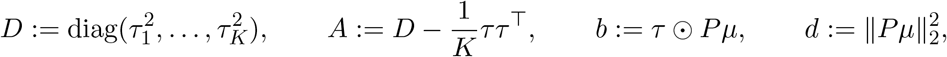

where ⊙ denotes entrywise product.

##### Lemma B.18

(Exact quadratic decomposition). *The sample variance satisfies*

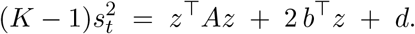

*Proof*. Use 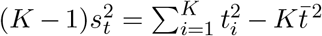, with 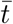 from (B.5), insert *t*_*i*_ = *τ*_*i*_*z*_*i*_ + *µ*_*i*_, and collect the quadratic, linear, and constant terms in *z*.

##### Lemma B.19

(Conditional mean and variance). *Condition on the deviation blocks (equivalently on τ, µ). Then*

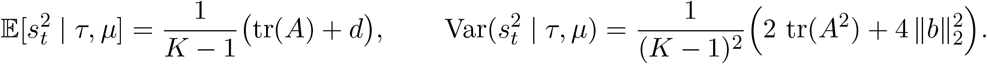

*Moreover, deterministically*,

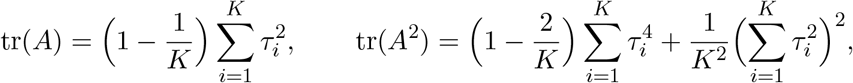

*and* 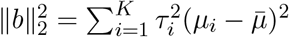 *with* 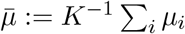.

*Proof*. By Lemma B.18 and *z* ∼ *N* (0, *I*_*K*_), use the Gaussian identities E[*z*^⊤^*Az*] = tr(*A*), Var(*z*^⊤^*Az*) = 2 tr(*A*^2^), E[*b*^⊤^*z*] = 0, 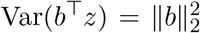, and Cov(*z*^⊤^*Az, b*^⊤^*z*) = 0. The closed forms follow by direct expansion of *A* and *b*.

##### Proposition B.20 (Upper bound on 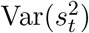 for *K* ≥ 10).

*Assume* (B.8) *with p* ∈ {1, 2, 4} *and K* ≥ 10. *Then, writing g* := *g*(*y*) *>* 0,

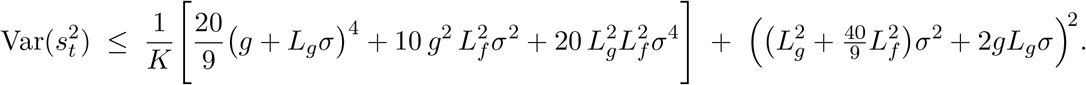

*Proof*. By the law of total variance,

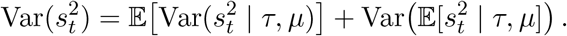

*Step 1: Gaussian-noise contribution*. From Lemma B.19,

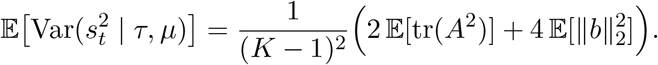

Using Lemma B.19 and 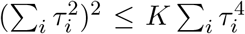 gives 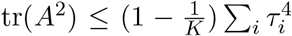. Next, let *a*_*i*_ := (E[|*δ*_*g,i*_|^4^])^1*/*4^. By Minkowski in ℝ^*K*^ with the *ℓ*_4_-norm,

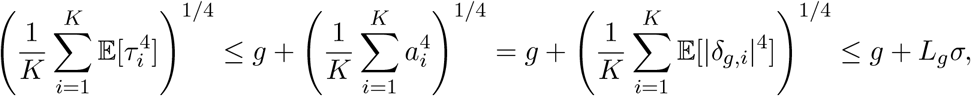

so 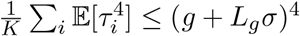, hence E[tr(*A*^2^)] ≤ (*K* − 1)(*g* + *L*_*g*_*σ*)^4^.

For 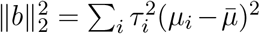, note 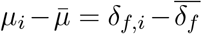. Using 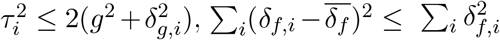, and Cauchy-Schwarz,

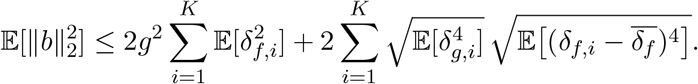

Moreover 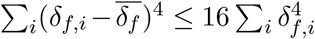 deterministically, hence 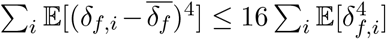. Applying Cauchy-Schwarz over *i* yields

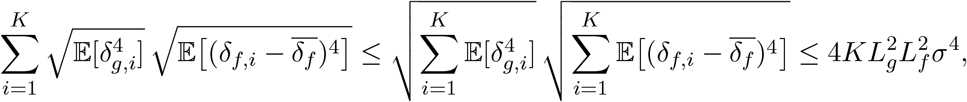

and 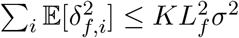 by (B.8) with *p* = 2. Therefore

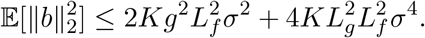

Substituting the bounds and using *K* ≥ 10 so that 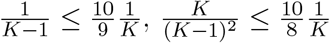 gives

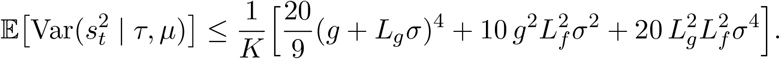

*Step 2: deviation-uncertainty contribution*. By Lemma B.19 and the identities therein,

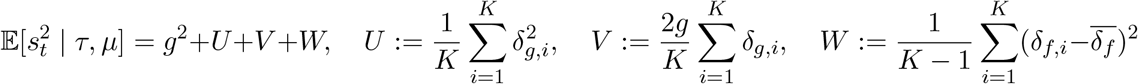

Using 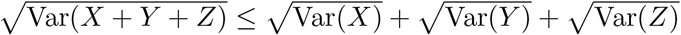,

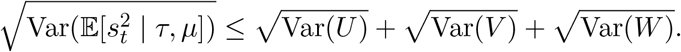

Without any independence across *i*, Var(*U*) ≤ E[*U* ^2^] and Cauchy-Schwarz imply

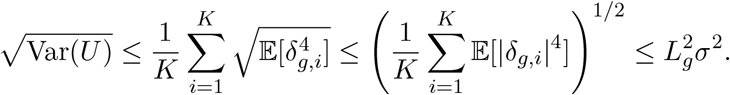

Similarly, 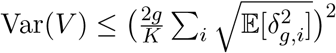 gives

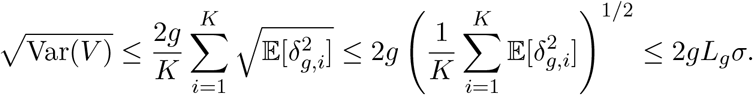

For *W*, use 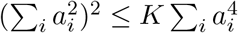 with 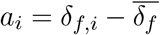 and 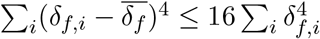 to obtain

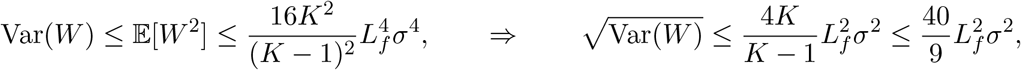

where the last inequality uses *K* ≥ 10. Therefore

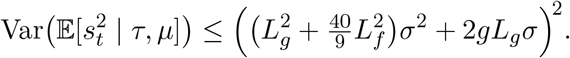

*Step 3: combine*. Summing the bounds from Steps 1–2 yields the claim.

##### Corollary B.21 (Simplified bound using *L* := max{*L*_*f*_, *L*_*g*_}).

*Under the assumptions of Proposition B.20, let L* := max{*L*_*f*_, *L*_*g*_}. *Then*

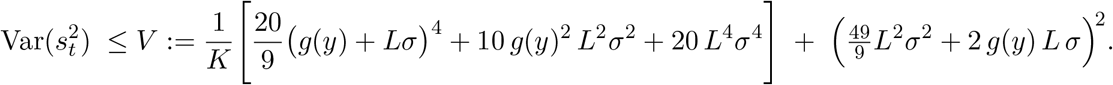

##### Corollary B.22 (Dimensionless bound)

*Under the assumptions of Corollary B.21, let L* := max{*L*_*f*_, *L*_*g*_}, *define* 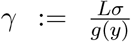, *and suppose ε* = *τ g*(*y*) *for some τ >* 0. *Since s*_*ε*_ ≥ *ε, we have g*(*y*) = *ε/τ. Then*

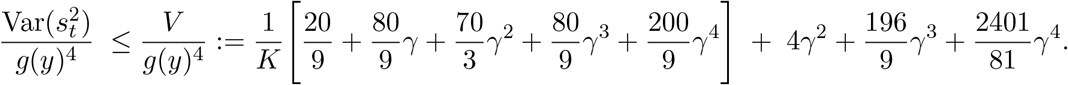

## Appendix C Properties of the ancestry representations *x* and *y*.

### C.1. Distributions of the ancestry representations

Ancestry adjustment of polygenic scores is typically performed using genetic ancestry representations such as Principal Component (PCA) projections or ancestry probabilities. Such representations are obtained by transforming genetic sequences into lower-dimensional representations, typically via linear projections, matrix factorization, or non-linear transformations.

PCA of genotype sequences has been shown to often correlate with the geographic location of populations and to capture genetic variation between ancestry groups [15, 7]. PCA provides a continuous representation that reflects population structures without relying on artificial ethnic-based categories. PCA has been one of the main covariates to try to remove non-genetic factors from GWAS and PRS development [14].

Unsupervised clustering for genotype sequences such as ADMIXTURE [1] or its modern variants [5, 19] predict a population percentage assignment for each sequence. Namely, each individual is assigned a point in the convex hull representing its population composition. Each cluster is then approximately matched to labels based on ethnic characteristics to aid interpretation.

Both PCA and clustering-based techniques provide bounded representations, as the genotype sequences are bounded and finite and the projection matrix used to obtain them are also bounded. It has been shown that all such representations follow an archetype-like structure, where a convex hull is formed, with some *founder* populations at the extremes and admixed populations in the interior [7]. Given this structure, it is sensible to assume that the ancestry follows a distribution that is compact and bounded as defined in the next section.

### C.2. The case *K* = *M* : averaging over all samples

*Setup*. Let *d* ≥ 1 denote the ambient dimension. Let 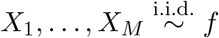 and let *Y* ∼ *f* be independent of 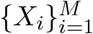. When emphasizing i.i.d. structure, we also write 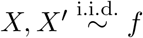 for two independent draws. Let *µ* := E[*X*] and Σ := Cov(*X*), so that tr(Σ) = E∥*X* − *µ*∥^2^. We also introduce the (dimensionless) radial fourth-moment ratio

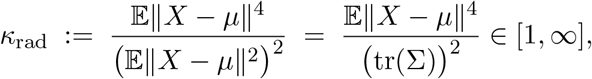

where the lower bound *κ*_rad_ ≥ 1 follows from Jensen’s inequality. When *K* = *M*, the ordered neighbors 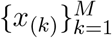 are simply a permutation of the samples 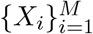, hence the averaged *p*th moment does not depend on sorting.

#### Lemma C.1 (Identity for *K* = *M*).

*For any p >* 0,

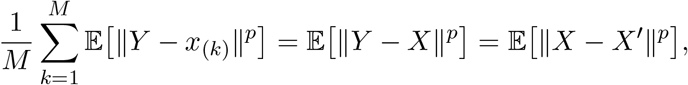

*where* 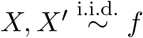 *are independent*.

*Proof*. Since 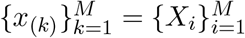 as multisets, 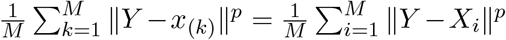 . Taking expectations and using i.i.d. gives 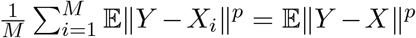 . Finally,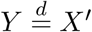 and *Y* ⊥ *X* implies 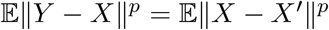.

*A generic bound for p* ∈ {1, 2, 4}. The cases *p* = 1, 2 depend only on tr(Σ), while *p* = 4 requires an additional tail/shape assumption. We adopt a bounded (vector) kurtosis condition, which is satisfied by many light-tailed families.

#### Proposition C.2 (A unified covariance-scale bound)

*Assume there exists κ*_4_ *<* ∞ *such that*

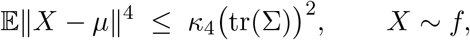

*namely κ*_rad_ ≤ *κ*_4_. *Define*

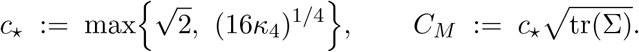

*Then, simultaneously for all p* ∈ {1, 2, 4},

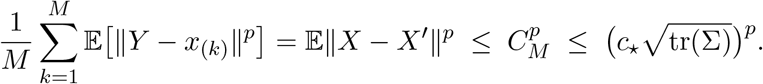

*Moreover, one may take the sharper (non-unified) constants*

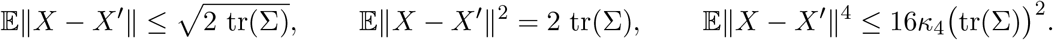

*Proof*. By Lemma C.1 it suffices to bound E∥*X* − *X*^′^∥^*p*^.

For *p* = 2, expand ∥*X* − *X*^′^∥^2^ = ∥(*X* − *µ*) − (*X*^′^ − *µ*)∥^2^ and use independence to obtain

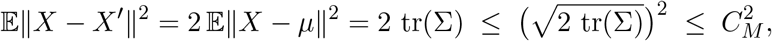

where the last inequality holds since 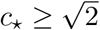.

For *p* = 1, Cauchy-Schwarz yields

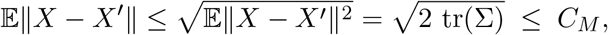

again because 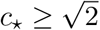.

For *p* = 4, use ∥*a* − *b*∥ ≤ ∥*a*∥ + ∥*b*∥ and (*u* + *v*)^4^ ≤ 8(*u*^4^ + *v*^4^) for *u, v* ≥ 0 to get

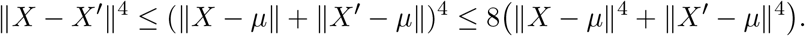

Taking expectations and using 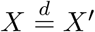 gives

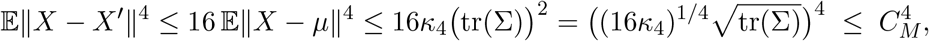

since *c*_⋆_ ≥ (16*κ*_4_)^1*/*4^.

Combining the above, for each *p* ∈ {1, 2, 4} we have 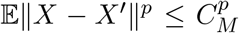 with the *same* constant 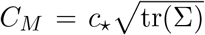, hence the bound holds simultaneously for all *p* ∈ {1, 2, 4}. The final inequality 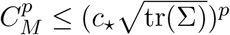 is immediate from the definition of *C*_*M*_ .

#### Corollary C.3 (A necessary lower bound for the unified constant)

*Under the assumptions and notation of Proposition C.2, any constant C*_*M*_ *satisfying*

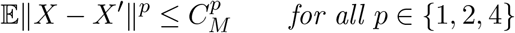

*must obey*

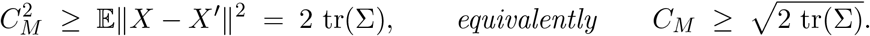

*In particular, for the choice* 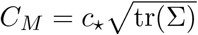 *in Proposition C.2, one necessarily has* 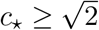.

*Examples: subgaussianity and bounded radius*. We record two common sufficient conditions and express the resulting bounds in the same 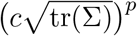 template.

#### Corollary C.4 Example: *c*-subgaussian implies bounded kurtosis)

*Assume Z* := *X* − *µ is c-subgaussian with covariance proxy* Σ, *i.e*.,

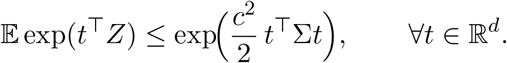

*Then there exists a universal constant C*_sg_ *(independent of d) such that*

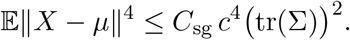

*Consequently, Proposition C.2 holds with*

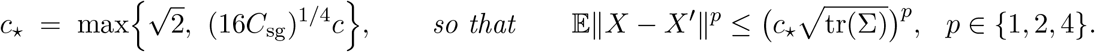

#### Corollary C.5 (Example: bounded radius expressed via tr(Σ))

*Assume* ∥*X* − *µ*∥ ≤ *R almost surely. Then for any p* ∈ {1, 2, 4},

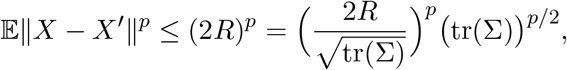

*(with the convention* 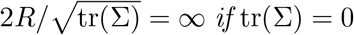 *if* tr(Σ) = 0 *and R >* 0*). In particular, Proposition C.2 holds with*

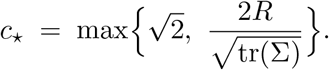

*Proof*. If ∥*X* − *µ*∥ ≤ *R* and ∥*X*^′^ − *µ*∥ ≤ *R*, then ∥*X* − *X*^′^∥ ≤ ∥*X* − *µ*∥ + ∥*X*^′^ − *µ*∥ ≤ 2*R* almost surely. The stated rewrite follows by factoring (tr Σ)^*p*/2^ .

### C.3. Moment bounds for *K*NN radii under bounded densities on a compact support

*Setup*. Let *S* ⊂ ℝ^*d*^ be compact with nonempty interior and diameter Δ := sup_*x,z*∈*S*_ ∥*x* − *z*∥ *<* ∞. Let *f* be a density supported on *S* with

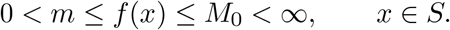

Assume a uniform interior-volume condition: there exists *κ* ∈ (0, 1] such that for all *y* ∈ *S* and *r* ∈ (0, Δ],

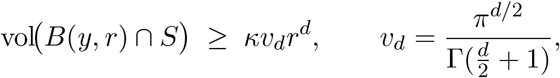

where vol(·) denotes *d*-dimensional Lebesgue volume and *B*(*y, r*) is the Euclidean ball. Let *X*_1_, …, *X*_*M*_ be i.i.d. from *f*, and let *y* ∈ *S* be independent of {*X*_*i*_}. Define *R*_*i*_ = ∥*X*_*i*_ − *y*∥ and order statistics *R*_(1)_ ≤ · · · ≤ *R*_(*M*)_. Let *x*_(*K*)_ achieve *R*_(*K*)_, so ∥*y* − *x*_(*K*)_∥ = *R*_(*K*)_.

*Geometric mass lower bound*. For *r* ∈ [0, Δ] define

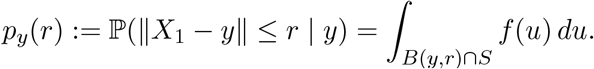

#### Lemma C.6

(Uniform ball-mass lower bound). *For all y* ∈ *S and r* ∈ [0, Δ],

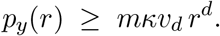

*Proof*. Using *f* ≥ *m* on *S* and the interior-volume condition, *p*_*y*_(*r*) ≥ *m* vol(*B*(*y, r*) ∩ *S*) ≥ *mκv*_*d*_*r*^*d*^ .

*A comparison coupling*. Set

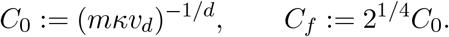

(The factor 2^1*/*4^ is only needed to cover the case *p* = 4 when *d* = 2, 3; see Lemma C.8.)

#### Lemma C.7 (Quantile comparison for *K*NN radii)

*Let* 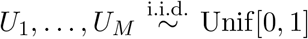 *and let W*_(*K*)_ *be the K-th order statistic. There exists a coupling such that, almost surely*,

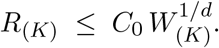

*Consequently, for any p >* 0,

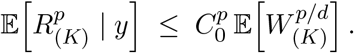

*Proof*. Lemma C.6 implies the conditional CDF *F*_*y*_(*r*) = ℙ(*R*_1_ ≤ *r* | *y*) satisfies *F*_*y*_(*r*) ≥ min{(*r/C*_0_)^*d*^, 1} for all *r* ≥ 0. A standard quantile coupling gives 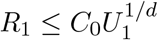 a.s.; taking *M* i.i.d. copies preserves coordinatewise domination and hence also order-statistic domination, yielding 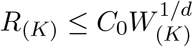. The moment bound follows by monotonicity.

#### Lemma C.8 (Moments of uniform order statistics)

*Let W*_(*K*)_ *be the K-th order statistic of M i.i.d*. Unif[0, 1] *variables. Then* E[*W*_(*K*)_] = *K/*(*M* + 1). *Moreover, for any a* ∈ (0, 2],

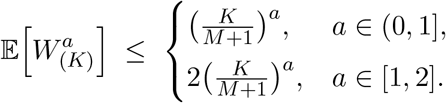

*Proof*. The identity E[*W*_(*K*)_] = *K/*(*M* + 1) is standard (e.g., by exchangeability of spacings). For *a* ∈ (0, 1], concavity of *t* ↦ *t*^*a*^ on [0, 1] gives 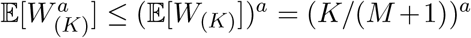. For *a* ∈ [1, 2], one may use the classical Gamma-ratio representation

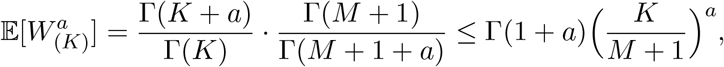

together with Γ(1 + *a*) ≤ Γ(3) = 2 on *a* ∈ [1, 2].

*Main bounds*. Throughout the remainder of this section we assume *d* ≥ 2 and consider *p* ∈ {1, 2, 4}.

#### Proposition C.9 (*K*NN moment bound for *p* ∈ {1, 2, 4}).

*Assume d* ≥ 2 *and let p* ∈ {1, 2, 4}. *Then for every* 1 ≤ *K* ≤ *M*,

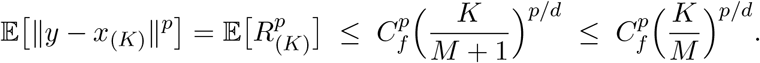

*Proof*. By Lemma C.7, 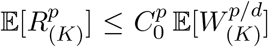. If *p* ∈ {1, 2} and *d* ≥ 2, then *p/d* ∈ (0, 1] and Lemma C.8 yields 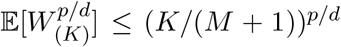, hence 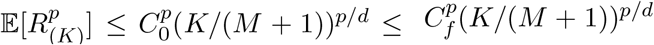. If *p* = 4, then 4*/d* ∈ (0, 2] for *d* ≥ 2, and Lemma C.8 gives 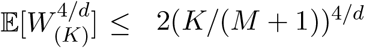, hence 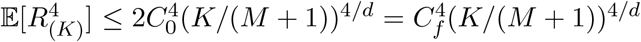. Finally, (*M* + _1)_^−*p/d*^ _≤ *M*_ ^−*p/d*^.

#### Proposition C.10 (Average moment bound over the first *K* neighbors)

*Assume d* ≥ 2 *and let p* ∈ {1, 2, 4}. *Then for every* 1 ≤ *K* ≤ *M*,

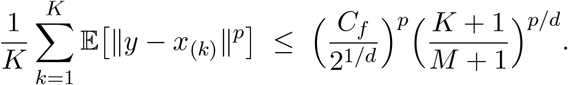

*Proof*. Apply Proposition C.9 with *K* replaced by *k* and average:

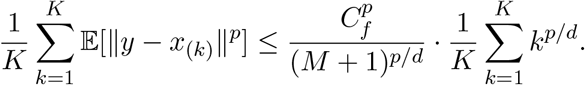

Since *p/d* ∈ (0, 1] for *p* ∈ {1, 2, 4} and *d* ≥ 2, the map *t* ↦ *t*^*p/d*^ is concave on [0, ∞), so

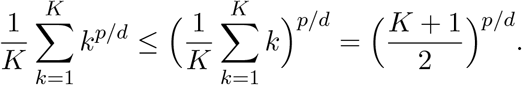

Rearranging yields the claim (this is exactly where the factor 2^−1*/d*^ can be absorbed into the constant).

#### Corollary C.11 (Clean *K/M* scaling)

*Assume d* ≥ 2 *and let p* ∈ {1, 2, 4}. *Then for every* 1 ≤ *K* ≤ *M*,

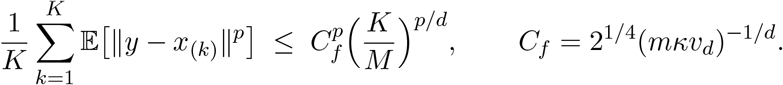

*Proof*. From Proposition C.10 it suffices to note that, for 1 ≤ *K* ≤ *M*,

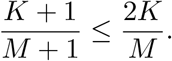

Plugging this into Proposition C.10 gives 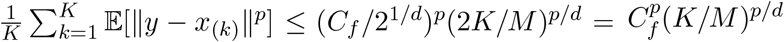.

#### Proposition C.12 (Optimal constant in the averaged *K/M* moment bound)

*Assume d* ≥ 2 *and fix p* ∈ {1, 2, 4}. *For all integers M* ≥ 1 *and* 1 ≤ *K* ≤ *M, define*

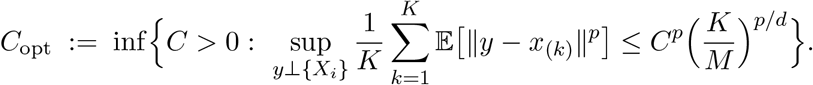

*Then C*opt *<* ∞ *and, under the standing assumptions*,

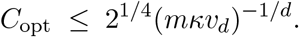

*Equivalently*,

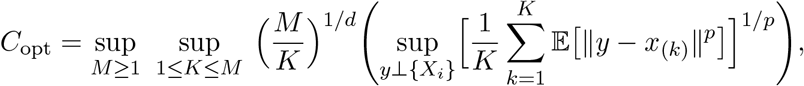

*and C*opt *may be approximated (for a fixed model class f and a chosen distribution of y) by numerical optimization over* (*M, K*) *together with Monte Carlo estimation of the averaged moment*.

*Proof*. The bound *C*opt ≤ 21/4(*mκvd*)−1*/d* follows from Corollary C.11. The supremum representation is obtained by rearranging the defining inequality and taking the smallest admissible *C*.

## Appendix D Mean Squared Error of 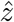.

### D.1. MSE decomposition for stabilized local standardization

Fix a target location *y* and recall the target draw *t* = *f* (*y*) + *g*(*y*)*z* from (B.3), with *z* ∼ *N* (0, 1). By iterated expectation,

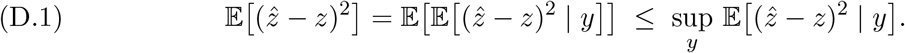

Define the stabilized local standardization estimator

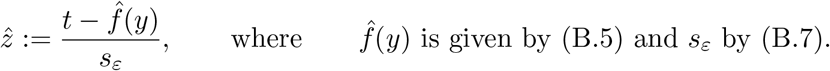

#### Lemma D.1 (Algebraic error identity)

*With t* = *f* (*y*) + *g*(*y*)*z and* 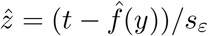,

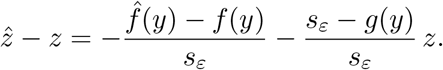

*Consequently*,

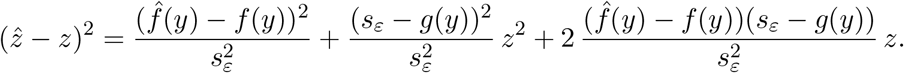

*Proof*. Since 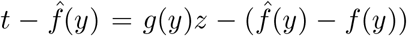, we have 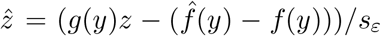. Subtract *z* and rewrite (*g*(*y*)*/sε* − 1)*z* = ((*g*(*y*) − *sε*)*/sε*)*z* to obtain the first identity; squaring yields the expansion.

#### Proposition D.2

(Exact conditional MSE decomposition). *Assume z is independent of the neighbor-based estimators* 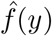 *and sε conditional on y. Then*

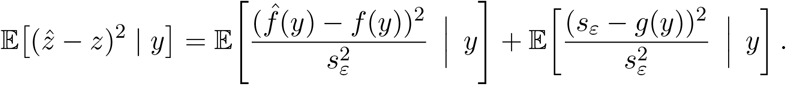

*Proof*. Take E[· | *y*] in Lemma D.1. Conditional independence implies E[*z*2 | *y*] = 1, and that 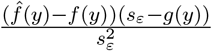 is independent of *z* given *y*, so the mixed term vanishes and the *z*2 term reduces to a factor 1.

#### Proposition D.3

(Conditional stabilized upper bound). *Under the assumptions of Proposition D.2, the stabilization sε* ≥ *ε from* (B.7) *implies*

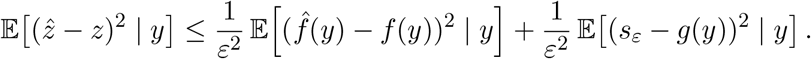

*Proof*. Apply Proposition D.2 and use the pointwise bound 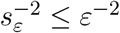, which follows from (B.7).

### D.2. Upper bounding 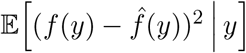

We bound the (unnormalized) conditional MSE of the local shift estimator 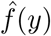 in terms of its conditional bias and variance.

#### Proposition D.4

(Conditional MSE via bias-variance). *For any estimator* 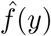 *measurable with respect to the neighbor block*,

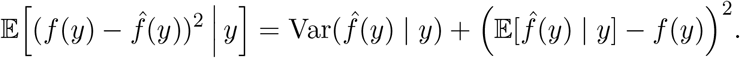

*Proof*. Expand around the conditional mean:

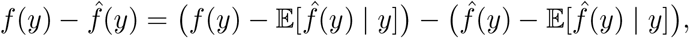

square, and take E[· | *y*]. The cross term vanishes because 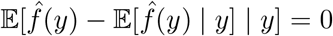.

#### Proposition D.5

(Moment upper bound for the shift MSE). *Assume* (B.8) *with p* = 1 *and p* = 2. *Then*

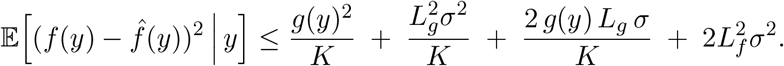

*Proof*. Combine Proposition D.4 with Proposition B.7 to upper bound 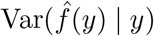, and with Corollary B.3 to upper bound the squared bias term 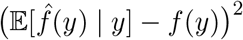 .

#### Corollary D.6

(Simplified normalized shift MSE). *Under the assumptions of Proposition D.5, let L* := max{*Lf, Lg*}, *define* 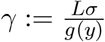, *and suppose ε* = *τ g*(*y*) *for some τ >* 0. *Since sε* ≥ *ε, we have*

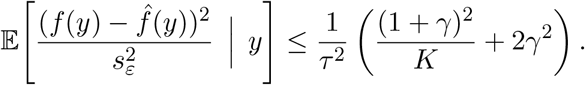

### D.3. Upper bounding 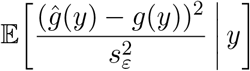

Fix a target location *y* and write *g* := *g*(*y*) *>* 0. Recall 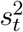 and *ĝ*(*y*) = *sε* from (B.6)–(B.7), where *sε* := max{*st, ε*} and *ε >* 0 is deterministic. Since *ĝ*(*y*) = *sε*, the normalized scale error is

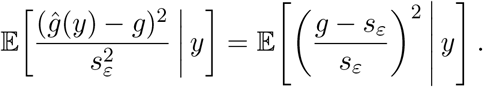

Throughout this subsection we assume conditional moment information for 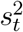:

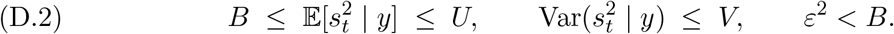

#### Lemma D.7

(Difference-of-squares identity). *For any s >* 0 *and g >* 0,

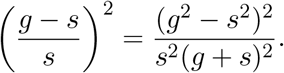

*Proof*. Use *g* − *s* = (*g*2 − *s*2)*/*(*g* + *s*) and square after dividing by *s*.

#### Lemma D.8

(Deterministic denominator control). *With sε* = max{*st, ε*} *and g >* 0,

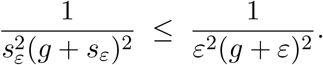

*Consequently*,

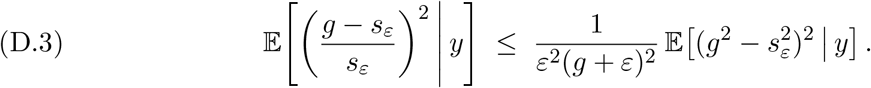

*Proof*. Since *sε* ≥ *ε*, we have 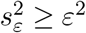 and *g* + *sε* ≥ *g* + *ε*. Taking reciprocals gives the first claim. Combine it with Lemma D.7 and take E[· | *y*] to obtain (D.3).

#### Lemma D.9

(Truncation split). *With sε* = max{*st, ε*},

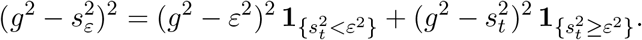

*In particular*,

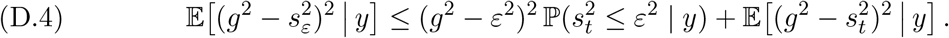

*Proof*. On {*st < ε*} we have *sε* = *ε*, and on {*st* ≥ *ε*} we have *sε* = *st*, which gives the identity. The inequality (D.4) follows by taking E[· | *y*] and dropping the indicator in the second term.

#### Lemma D.10

(Cantelli bound for the small-scale event). *Under* (D.2),

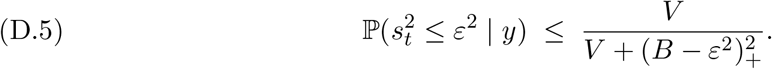

*Proof*. Let 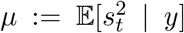 and set *a* := (*µ* − *ε*2)+ ≥ 0. Cantelli’s inequality applied conditionally to 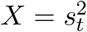 yields

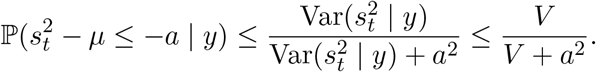

Since *µ* ≥ *B*, we have (*µ* − *ε*2)+ ≥ (*B* − *ε*2)+, hence 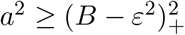, which gives (D.5).

#### Lemma D.11

(Second-moment control of 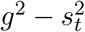). *Under* (D.2),

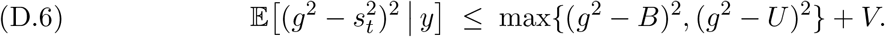

*Proof*. Let 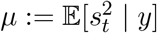. Decompose 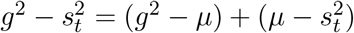 and expand:

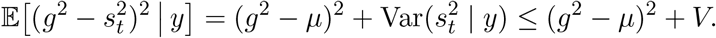

Since *µ* ∈ [*B, U* ] and *x* 1→ (*g*2 − *x*)2 is convex, (*g*2 − *µ*)2 ≤ max{(*g*2 − *B*)2, (*g*2 − *U*)2} gives (D.6).

#### Proposition D.12

(General bound from moments of 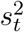). *Assume* (D.2). *Then*

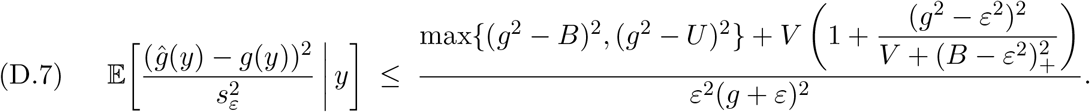

*Proof*. By Lemma D.8, it suffices to upper bound 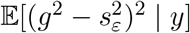. Lemma D.9 gives the reduction (D.4). Bound the probability term using Lemma D.10, and bound 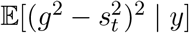 using Lemma D.11. Substitute the resulting bound into (D.3) to obtain (D.7).

#### Lemma D.13

(Uniform *<* 2 envelope for the truncated-scale ratio). *Fix y and write g* := *g*(*y*) *>* 0. *Let ε* = *τg with* 0 *< τ <* 1, *and define* 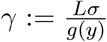 *with L* := max{*Lf, Lg*}. *Let V be as in Corollary B.22 and let B be as in Corollary B.11. Then, for every γ >* 0,

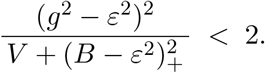

*Proof*. Set *a* := 1 − *τ* 2 ∈ (0, 1), so (*g*2 − *ε*2)2 = *a*2*g*4. By Corollary B.11,

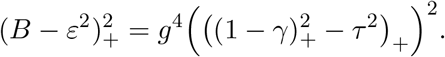

For *γ* ∈ [0, 1] we have (1 − *γ*)+ = 1 − *γ* and hence 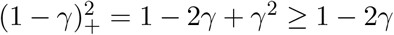, while for *γ >* 1 the left-hand side equals 0. Thus, for all *γ* ≥ 0,

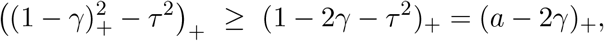

and consequently 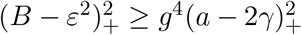.

Next, by Corollary B.22, dropping the nonnegative 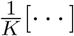 term yields

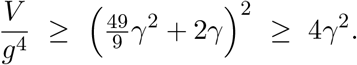

Combining the last two displays gives

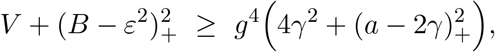

so

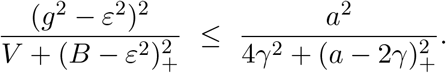

A direct minimization shows 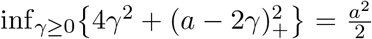, attained at *γ* = *a/*4 ∈ (0, *a/*2), whence the right-hand side is at most 2. Moreover, for every *γ >* 0 at least one of the previous lower bounds is strict, so the original ratio is *<* 2.

#### Lemma D.14

(Exact normalized max squared deviation). *Assume the hypotheses of Corollaries B.17 and B.11. Let g* := *g*(*y*) *>* 0 *and* 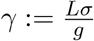. *If K* ≥ 10, *then*

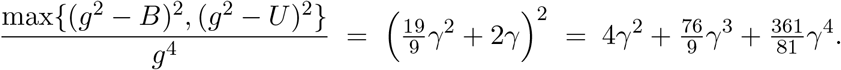

*Proof*. By Corollary B.17, 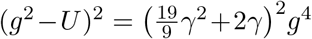. By Corollary B.11, 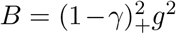, hence 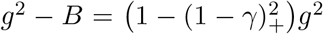, which equals (2*γ* − *γ*2)*g*2 if *γ <* 1 and equals *g*2 if *γ* ≥ 1. If *γ <* 1, then 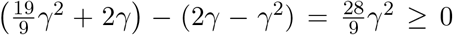, so *U* ™ *g*2 ≥ *g*2 – *B*; if *γ* ≥ 1, then 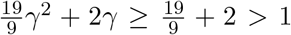 so again *U* − *g*2 ≥ *g*2 − *B*. Thus (*g*2 − *U*)2 ≥ (*g*2 − *B*)2 and the maximum equals (*g*2 − *U*)2; dividing by *g*4 gives the first equality, and the polynomial form follows by expanding.

#### Proposition D.15

(Polynomial MSE bound for truncated-scale normalization). *Assume* (D.2) *and the hypotheses of Corollary B.22, Corollary B.11, and Corollary B.17. Fix y and write g* := *g*(*y*) *>* 0. *Let ε* = *τg with* 0 *< τ <* 1, *define* 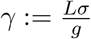 *with L* := max{*Lf, Lg*}, *and assume K* ≥ 10. *Then*

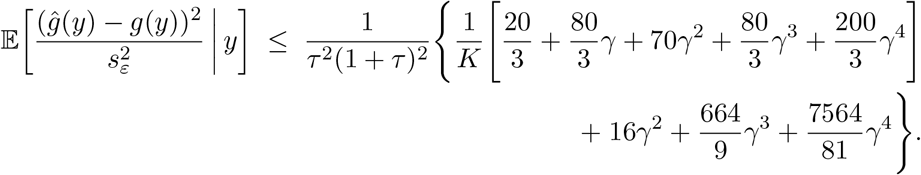

*Proof*. Apply Proposition D.12 and substitute *ε* = *τg*. Normalize by *g*4. Use Lemma D.13 to bound the truncated-scale ratio, giving the factor 1 + · *<* 3 on the *V/g*4 contribution. Use Lemma D.14 for the normalized max squared deviation term. Finally, invoke Corollary B.22 for *V/g*4, multiply its 1*/K* polynomial by 3, and add the max-deviation polynomial to obtain the displayed bound.

### D.4. Upper bounding 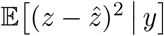

#### Proposition D.16

(Bound on the total normalized MSE). *Assume the hypotheses of Corollary D.6 and Proposition D.15. Fix y and write g* := *g*(*y*) *>* 0. *Let ε* = *τg with* 0 *< τ <* 1, *and define* 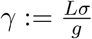 *with L* := max{*Lf, Lg*}. *If K* ≥ 10, *then*

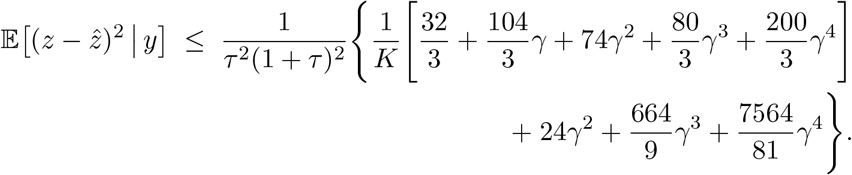

*Proof*. Using the decomposition from Proposition D.5, the conditional MSE splits into the normalized shift term and the normalized scale term as 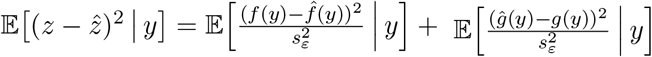. Bound the shift term by Corollary D.6, multiply by (1 + *τ*)2*/*(1 + *τ*)2, and use 0 *< τ <* 1 so that (1 + *τ*) *<* 2 to upper bound the resulting numerator factor by 4. Bound the scale term by Proposition D.15. Summing the two bounds and collecting the *K*−1 and *γ*-polynomial contributions yields the stated inequality.

#### Corollary D.17

(Quadratic bound under 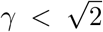). *Under the assumptions of Proposition D.16, suppose in addition that* 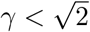. *Then, for K* ≥ 10,

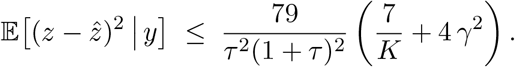

*Proof*. From Proposition D.16 and 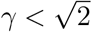 we have 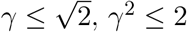, hence 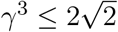 and *γ*4 ≤ 4, so 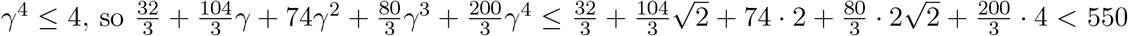 and 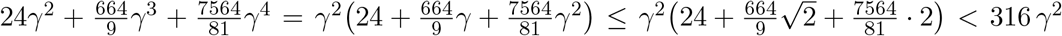. Plugging these into Proposition D.16 yields 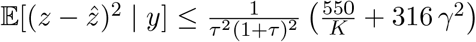. Finally, since 79 · 7 = 553 ≥ 550 and 79 · 4 = 316, we have 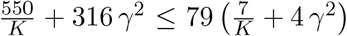, concluding the proof.

### D.5. Upper bounding 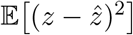

#### Proposition D.18

(Quadratic bound with *K/M* -controlled *γ*). *Under the assumptions of Proposition D.16 and Corollary D.17, assume in addition that d* ≥ 2, *the covariate y* ∼ *p is sampled independently of the reference set* 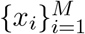, *the sampling density p satisfies the standing assumptions of Section C.3 (so Corollary C.11 applies), and that gℓ* ≤ *g*(*y*) *for all y* ∈ ℝ*d. Let τ* ∈ (0, 1) *and define* 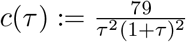.

*Let* 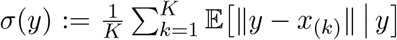 *and* 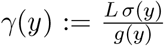, *and define* 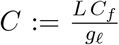 *is as in Corollary C.11. If K* ≥ 10 *and* 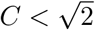, *then*

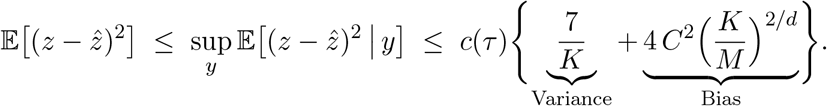

*Proof*. Since 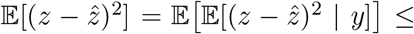 sup*y* 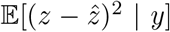, it suffices to bound 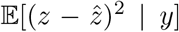 uniformly in *y*. By Corollary C.11 with 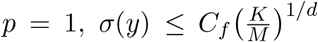, hence 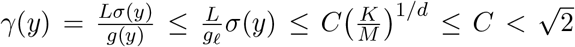. Therefore Corollary D.17 applies for all *y* (using *K* ≥ 10), giving 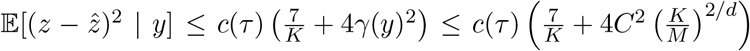, and taking sup*y* yields the claim.

## Acknowledgments

We would like to acknowledge the helpful discussions on the ancestry adjustment problem with Elisabeth Rosenthal, and thank the other members of the Ioannidis Lab for useful feedback on the manuscript. This work uses data provided by patients and collected by the NHS as part of their care and support. We thank the UK Biobank and its participants and the NHS. This research has been conducted using the UK Biobank Resource under Application Number 89006. This work was supported in part by NHGRI grant number HG013628 awarded to AGI and CDB.

## Author Contributions

All authors have made substantial intellectual contributions to the study conception, execution, and design of the work. All authors have read and approved the final manuscript. In addition the following contributions are noted: Conceptualization: DMM, MB, CDB, AGI; Methodology: DMM, MB; Formal analysis and investigation: DMM; Writing - original draft preparation: DMM, MB, CDB, AGI; Writing - review and editing: DMM, MB, CDB, AGI; Funding acquisition: CDB, AGI; Resources: DMM, MB, CDB, AGI; Supervision: CDB, AGI.

